# Models and modelling practices for assessing the impact of outbreak response interventions to human vaccine-preventable diseases (1970-2019) - A systematic review

**DOI:** 10.1101/2022.05.27.22275642

**Authors:** James M. Azam, Xiaoxi Pang, Elisha B. Are, Juliet R.C. Pulliam, Matthew J. Ferrari

## Abstract

**Background:** Mathematical modelling can aid outbreak response decision-making. However, this would require collaboration among model developers, decision-makers, and local experts to incorporate appropriate realism. We conducted a systematic review of modelling studies on human vaccine-preventable disease (VPD) outbreaks to identify patterns in modelling practices among collaborations. We complemented this with a mini review of eligible studies from the foot-and-mouth disease (FMD) literature.

**Methods:** Three databases were searched for studies published during 1970-2019 that applied models to assess the impact of an outbreak response. Per included study, we extracted data on author affiliation type (academic institution, governmental, and non-governmental organizations), whether at least one author was affiliated to the country studied, interventions, and model characteristics. Furthermore, the studies were grouped into two collaboration types: purely academic (papers with only academic affiliations), and mixed (all other combinations) to help investigate differences in modelling patterns between collaboration types in the human disease literature. Additionally, we compared modelling practices between the human VPD and FMD literature.

**Results:** Human VPDs formed 228 of 253 included studies. Purely academic collaborations dominated the human disease studies (56%). Notably, mixed collaborations increased in the last seven years (2013 - 2019). Most studies had an author in the country studied (75.2%) but this was more likely among the mixed collaborations. Contrasted to the human VPDs, mixed collaborations dominated the FMD literature (56%). Furthermore, FMD studies more often had an author affiliated to the country studied (92%) and used complex model design, including stochasticity, and model parametrization and validation.

**Conclusion:** The increase in mixed collaboration studies over the past seven years could suggest an increase in the uptake of modelling for outbreak response decision-making. We encourage more mixed collaborations between academic and non-academic institutions and the involvement of locally affiliated authors to help ensure that the studies suit local contexts.

## Introduction

Successful outbreak response to infectious diseases is often the result of a highly collaborative process [1]. Decision-making during outbreak response usually requires collaborations between academic and field experts, including governmental and Non-Governmental Organizations (NGOs) [1–5]. These collaborations ensure that important perspectives from both research and operations/implementation are accounted for in the decision-making process.

Outbreak response decision-making often requires adaptations to the affected location [1, 5, 6]. The interventions and decisions are usually driven by data/evidence, information, and experiences from the past or in real-time, either locally or from similar phenomena elsewhere [3, 7]. Furthermore, involving local experts in the design of interventions has been found to boost the success of outbreak response efforts [1]. Consequently, outbreak response teams should ideally involve at least one local expert to provide more context [1].

Mathematical modelling can support outbreak response decision-making [8]. Modelling is a proven tool for revealing insights about the extent of disease spread, and impact of interventions, while drawing on lessons learnt to provide recommendations for decision-making during outbreaks [2, 5, 8]. Insights from mathematical modelling, though often useful, only form part of the larger context (socio-economic, political, and so forth) to be considered during an outbreak, making it difficult to determine the extent to which it contributes to outbreak response decision-making [2, 3]. One way to help ensure that modelling contributes to decision-making could be through the conduct of interdisciplinary research between model developers and decision-makers.

The link between interdisciplinarity in scientific research — that is, research conducted by authors with diverse scientific backgrounds — and research impact, for example, number of citations has been well-researched [9, 10]. However, few studies have investigated the impact of interdisciplinary collaborations on the conduct of scientific research. One such study investigated the impact of interdisciplinarity on the scientific validity of the methods used in a selection of papers that applied machine learning on topics in biology or medicine [11]. The study found that the methods used in the reviewed studies differed according to the nature of the scientific backgrounds of the authors who conducted the research. Our preliminary searches of the literature did not reveal any of such review studies investigating differences in research practices among collaborations in the outbreak response modelling literature. We, therefore, conducted a systematic review of outbreak response modelling studies of human vaccine-preventable diseases with the goal to explore the time and geographic (countries) patterns of collaboration types, and to ascertain differences in practices between the collaboration types in terms of the choice of model characteristics, modelling methods, and other topics.

We sought to answer the following questions with respect to the collaboration landscape in the human disease outbreak response modelling literature: (1) How are the studies distributed in time and geographic locations, and how often is at least one of the collaborators geographically connected to the location being studied? (2) What types of interventions are generally modelled and is there a difference between the conclusions drawn about the impact of interventions, especially, vaccination in comparison with other non-vaccination interventions during outbreaks? (3) Do the model characteristics (structure and dynamics) and modelling practices (parametrization, validation, and so forth) differ between the collaboration types?

We acknowledge that there is a parallel body of relevant modelling work in the livestock literature. Specifically, the outbreak response effort that resulted from the 2001 foot-and-mouth disease (FMD) virus outbreak in the United Kingdom has been a foundational example of the application of mathematical modelling for decision-making. Subsequent work based on this outbreak has been instrumental in the development of FMD outbreak intervention strategies around the world. Hence, even though the primary scope of this review is the application of models to inform outbreak response for human infectious diseases, we include the relevant FMD references for comparison. In the end, we, therefore, study the differences in terms of collaborations in the human disease modelling literature, and compare our observations to that of the FMD modelling literature to further ascertain any differences in approaches and practices between the human and livestock literature.

## Materials and methods

We followed the 2020 Preferred Reporting Items for Systematic Reviews and Meta-analyses statement (PRISMA 2020) to conduct this systematic review [12].

### Eligibility criteria

The following definitions were used in the determining study eligibility:

- “Outbreak”: a new and sudden rise in the number of cases of a disease in a population, which when left uncontrolled, could lead to large scale geographic spread, AND
- “Outbreak response”: an intervention directly triggered by the outbreak of an infectious disease, AND
- “Mechanistic models”: mathematical models that use an equation or system of equations to capture the biological mechanisms driving the transmission dynamics of the infectious disease at the level of an individual or population [13, 14], AND
- “Outbreak intervention assessment”: a mechanistic model-based evaluation of the impact of an outbreak response intervention.

We included studies that used a mechanistic model to investigate the impact of interventions triggered by the outbreak of any of the human vaccine-preventable diseases we considered (Table 1 in S1 File), Ebola, or foot-and-mouth disease. Studies were considered unique even if they had a duplicated author list or a slight variation in the author list, probably representing the same modelling group, so far as the content of the paper was different.

**Table 1.** Number of studies per unique combination of author affiliations (human diseases). Here, we also show the grouping of author affiliation combinations into purely academic and mixed collaborations. Purely academic collaborations refer to those authored by only authors with academic institution affiliations whereas mixed collaborations include those authored by a mixture of authors with academic institution affiliations, government institutions, and NGOs or only one of the last two.

| Collaboration type | Author affiliation combination | Number of publications |
| --- | --- | --- |
| Purely academic | academic only | 128 |
| Mixed | academic + governmental | 57 |
| Mixed | academic + NGO | 17 |
| Mixed | academic + governmental + NGO | 14 |
| Mixed | governmental only | 5 |
| Mixed | NGO only | 5 |
| Mixed | governmental + NGO | 2 |
| **Total** |  | **228** |

We excluded studies that satisfied at least one of the following criteria:

- Study used a model that is not mechanistic,
- Study is not about an outbreak as defined,
- Disease studied is not a human vaccine-preventable disease (Table 1 in S1 File), Ebola or foot-and-mouth disease,
- Study’s objective is not to evaluate the impact of an intervention mounted in response to an outbreak, or
- Study is not published in English.

### Information sources

On January 15, 2020, we searched Scopus, PubMed, and Web of Science for eligible records. We searched each database from its earliest date of coverage through January 15, 2020.

### Search strategy

We constructed and validated search strings specific to each database. To validate the search strings, we first ran them in the specific database’s search engine and obtained a database of records. We then searched the database for well-known papers that fit the criteria for included studies. We did this for each disease on our list.

The search strings were constructed by all five reviewers (JMA, XP, EBA, MJF, and JRCP) and in consultation with the Faculty of Science Librarian of Stellenbosch University to reflect three main topics and their synonyms, that is, “outbreak”, “intervention”, and “mechanistic model”.

The following are the search strings per database.

#### Scopus (Title, abstract, keywords search)

(TITLE-ABS-KEY (epidemic OR outbreak OR emergency OR reactive OR crisis)) AND (TITLE-ABS-KEY (respon* OR manage* OR control OR interven* OR strateg*)) AND (TITLE-ABS-KEY (stochastic OR transmission OR computational OR mathematical OR mechanistic OR statistical OR simulation OR “In silico” OR dynamic*)) AND (TITLE-ABS-KEY (model*)) AND ((TITLE-ABS-KEY (cholera OR dengue OR diphtheria OR ebola OR “Foot-and-mouth” OR “foot and mouth” OR fmd OR “Hepatitis A” OR “Hepatitis B” OR “Hepatitis E” OR “Haemophilus influenzae type b” OR hib OR “Human papillomavirus” OR hpv OR influenza)) OR (TITLE-ABS-KEY (“Japanese encephalitis” OR malaria OR measles OR “Meningococcal meningitis” OR mumps OR pertussis OR “Whooping cough” OR “Pneumococcal disease” OR poliomyelitis OR polio OR rabies OR rotavirus OR rubella)) OR (TITLE-ABS-KEY (tetanus OR “Tick-borne encephalitis” OR tuberculosis OR typhoid OR varicella OR chickenpox OR “Yellow Fever” OR “vaccine-preventable”)))

#### PubMed (Title and abstract search)

Search ((((((Epidemic OR Outbreak OR Emergency OR Reactive OR Crisis))) AND ((Response OR Management OR Control OR Intervention OR Strategies))) AND ((Stochastic OR Transmission OR Computational OR Mathematical OR Mechanistic OR Statistical OR Simulation OR “In silico” OR Dynamic*))) AND model*) AND ((Cholera OR Dengue OR Diphtheria OR Ebola OR “Foot-and-mouth” OR “foot and mouth” OR FMD OR “Hepatitis A” OR “Hepatitis B” OR “Hepatitis E” OR “Haemophilus influenzae type b” OR Hib OR “Human papillomavirus” OR HPV OR Influenza OR “Japanese encephalitis” OR Malaria OR Measles OR “Meningococcal meningitis” OR Mumps OR Pertussis OR “Whooping cough” OR “Pneumococcal disease” OR Poliomyelitis OR Polio OR Rabies OR Rotavirus OR Rubella OR Tetanus OR “Tick-borne encephalitis” OR Tuberculosis OR Typhoid OR Varicella OR Chickenpox OR “Yellow Fever” OR “vaccine-preventable”))

#### Web of Science (Topic search)

TOPIC: (Epidemic OR Outbreak OR Emergency OR Reactive OR Crisis) AND TOPIC: (Respon* OR Manage* OR Control OR Interven* OR Strateg*)) AND TOPIC: (Stochastic OR Transmission OR Computational OR Mathematical OR Mechanistic OR Statistical OR Simulation OR In silico OR Dynamic*) AND TOPIC: (model*) AND TOPIC: (Cholera OR Dengue OR Diphtheria OR Ebola OR “Foot-and-mouth” OR “foot and mouth” OR FMD OR “Hepatitis A” OR “Hepatitis B” OR “Hepatitis E” OR “Haemophilus influenzae type b” OR Hib OR “Human papillomavirus” OR HPV OR Influenza OR “Japanese encephalitis” OR Malaria OR Measles OR “Meningococcal meningitis” OR Mumps OR Pertussis OR “Whooping cough” OR “Pneumococcal disease” OR Poliomyelitis OR Polio OR Rabies OR Rotavirus OR Rubella OR Tetanus OR “Tick-borne encephalitis” OR Tuberculosis OR Typhoid OR Varicella OR Chickenpox OR “Yellow Fever” OR “vaccine-preventable”)

### Study selection

We used Endnote version X7.8 and Rayyan web application (https://www.rayyan.ai/) to combine the results from the three databases and to identify and remove duplicate records. The unique records were exported into Rayyan web application, where they were screened in two stages by three reviewers (JMA, XP, and EBA).

Stage one screening involved excluding studies based on their title and abstract, using the questionnaire that follows. The reviewers only used the information provided in the title and abstract of each study to decide whether it was eligible for inclusion (“Include”), not eligible (“Exclude”), or likely to be included (“Maybe”). The “Include” and “Maybe” category of studies further went through stage two screening described ahead, which was more stringent.

#### Questionnaire for title and abstract screening (Stage 1)

1. Is this article written in English?
  - No: Exclude. Reason: Not English
2. Does the title of this article fit the scope of this review?
  - No: Exclude. Reason: Title out of scope
3. Does the topic of the abstract fit the scope of this review?
  - No: Exclude. Reason: Topic out of scope
4. Is this study entirely a review (literature review, systematic review, scoping review, not indicated)?
  - Yes: Exclude. Reason: Review
5. Is any part of this study about an outbreak that happened in the past, was ongoing at the time of the study, or a hypothetical one, including one that could happen in the future? We define an outbreak as a new and sudden rise in the number of cases of a disease in a population, which when left uncontrolled, could lead to large scale geographic spread.
  - No: Exclude. Reason: Not an outbreak
6. Is any part of this study about a human infectious disease (Table 1 in S1 File), Ebola or foot-and-mouth disease in livestock?
  - No: Exclude. Reason: Disease not in scope
7. Does this study assess the impact – epidemiological or operational - of a real or hypothetical intervention that was mounted or could potentially be mounted in response to an outbreak of the disease in question? Note that we define an assessment as an evaluation of either the absolute or relative impact of an intervention on one of several outcomes including number/proportion of population reached with the intervention (coverage), and a change in size - number of people, duration, spatial - of the outbreak.
  - No: Exclude. Reason: Not an outbreak intervention assessment
  - Maybe. Reason: Intervention details unclear
8. Does this study solely use static methods for evaluating the intervention? Static methods include surveys, regression methods, descriptive, and exploratory statistical methods.
  - No: Exclude. Reason: Not a model
9. Does this study use any kind of equation, system of equations, or computer simulation to capture the disease’s transmission process or natural history over time?
  - No: Exclude. Reason: Model not mechanistic
  - Maybe. Reason: Likely a model

Stage one was first piloted with all three reviewers screening the same 50 studies using Rayyan in “blind mode”. This was so that the reviewers could not see each other’s screening decisions (inclusion/exclusion/maybe). After the pilot screening, the results were compared and any inclusion/exclusion conflicts were discussed and resolved. The pilot phase ensured that all the reviewers were using a consistent screening approach. The remaining studies were then screened in duplicate and all conflicting decisions were resolved at the end through discussions among the reviewers.

Stage 2 involved screening the included studies from stage 1, using their full text, in duplicate and with the following questionnaire.

#### Questionnaire for full text screening (Stage 2)

1. Is this article written in English? (This was necessary because some articles could have English abstracts but non-English full text)
  - If no, exclude. Reason: Not English
2. Is this article a report, commentary or any kind of non-quantitative report?
  - If yes, exclude. Reason: Article type out of scope
3. Is the full text readily available?
  - If no, mark as Maybe. Reason: Full text not available
4. Is this study entirely a review (literature review, systematic review, scoping review, not indicated)?
  - If yes, exclude. Reason: Review
5. Is the study or a part of it about an outbreak, real or hypothetical?
  - If no, exclude. Reason: Not an outbreak
6. Is the disease a human vaccine-preventable disease (Table 1 in S1 File), Ebola, or foot-and-mouth disease?
  - If no, exclude. Reason: Not a listed disease
7. Does this study solely use static methods for evaluating the intervention? Static methods include surveys, regression methods, descriptive, exploratory statistical methods.
  - If no, exclude. Reason: Static model or method
8. Does this study use any kind of equation, system of equations, or computer simulation to capture the disease’s transmission process in a dynamic way?
  - If no, exclude. Reason: Model not mechanistic
  - If unclear, maybe. Reason: Likely a model
9. Is the model about within-host dynamics?
  - If yes, exclude. Reason: Within-host model
10. Does this study attempt to assess the impact – epidemiological or operational - of a real or hypothetical intervention that was mounted or could potentially be mounted in response to an outbreak of the disease in question? We define an assessment as an evaluation of either the absolute or relative impact of an intervention on one of several outcomes including number/proportion of population reached with the intervention (coverage), and a change in size - number of people, duration, spatial - of the outbreak.
  - If no, exclude. Reason: Not an outbreak intervention assessment
  - If unclear, maybe. Label: Intervention details unclear

### Data extraction

To extract the data from the studies, we used the data extraction questionnaire implemented with the KoboToolbox web application (Section 3 in S1 File). The questionnaire was first piloted with 10 papers among the three reviewers, who worked in duplicate. All conflicts in terms of extracted data were resolved.

Following the pilot phase, the full list of included studies was shared among three reviewers (JMA, XP, and EBA), who extracted the relevant data independently. The extracted data was cross-checked by one reviewer (JMA). When a discrepancy was found in the extracted data, the reviewer referred to the original paper and resolved the conflict.

We extracted data on the type of publication, author affiliation type (academic/government/non-governmental), country/setting studied, whether at least one author affiliation was situated in the country studied, disease studied, the study objectives (retrospective/prospective impact assessment, timing of the modelling practice with respect to the outbreak (retrospective/prospective/real-time), interventions studied, and whether vaccination was the most impactful intervention when compared as a single intervention (yes/no/inconclusive). A full list of the extracted data items are provided (Section 2 in S1 File).

From the model in each paper, we extracted data on the representation of individuals (compartmental vs individual-based), whether spatial structure was represented (yes/no), model structure (deterministic vs stochastic), model parameterization, and validation. We also extracted a predefined list of outcomes (cases averted, final epidemic size, etc) measured with the models. If an outcome was reported that was not on the pre-defined list, we collected it separately as free text for further analysis. We also extracted data on whether the studies included sensitivity analysis or not. This was to understand how uncertainties in the model inputs were dealt with. We collected data on whether the studies used real world data, that is, excluding simulated data, and whether the data was openly available for download. We also extracted information on whether the code used for the analysis and visualization were made openly available.

### Data analysis and synthesis

To understand how the papers were distributed in terms of author affiliations, we first tallied the unique combinations of author affiliation types. We then grouped the author affiliations into the following two collaboration types and performed all further analyses by stratifying the variable(s) of interest by them:

1. *Purely academic:* papers with only academic author affiliations (academics collaborating with other academics),
2. *Mixed:* papers with a mixture of academic, government and NGO affiliations. This collaboration type also included papers with only government or only NGO affiliations.

We first explored the trend in aggregated publications during 1970-2019 by counting the number of publications in each year irrespective of collaboration type.

To explore how the publications changed in time with respect to the collaboration types, we tallied the number of publications and relative proportion (mixed versus purely academic collaboration) of studies per year. We lumped together the publications from 1970 to 2005 due to the small numbers.

To study how the papers were distributed in terms of geographic locations, we split the studies into those that studied actual versus hypothetical locations and used the subset of studies about actual locations to rank the topmost studied locations (country and continent) by frequency.

We also investigated how connected the authors were to the locations studied. Here, we only considered studies on geographic scales up to actual countries and not larger. All studies about locations larger than a country were dropped from this part of the analysis. We tallied, in terms of collaboration types, the number of studies about actual locations (countries) with or without at least one author affiliation in the studied location. If a paper studied more than one location, we treated each location as a separate instance. This increased the denominators accordingly.

To summarize the conclusions drawn about the impact of vaccination as a single intervention, we grouped the studies into three categories, that is, studies that modelled non-vaccination interventions, those that modelled vaccination in combination with other interventions, and those that modelled vaccination as a single intervention for comparison with other interventions. The question was only applicable to the last category of interventions, so we counted the number of studies that found vaccination to be the most impactful or not. Where vaccination was not found to be the most impactful, we briefly summarised the reasons and modelling assumptions.

For the disease studied, model characteristics (structure, dynamics, and spatial heterogeneity), and modelling practices (parametrization, validation, sensitivity analysis, data and code use and availability), we counted the number of studies per collaboration type and reported the results as percentages and fractions of the total. In counting the number of papers per disease studied and outcome measured, if a study had more than one outcome measured or disease, we treated each instance as unique and counted them separately. Hence, in such cases, the denominator of the reported fraction increased from the total studies we reported in our search results.

To summarise the model outcomes, we stratified by the collaboration type and tallied the outcomes used within each group. We reported the top six most used outcomes by both collaboration types and compared the results between the two.

All analyses were perform in R 4.0.5 [15]. We have provided a database of all the screened papers and their associated extracted data in a csv file, which can be accessed at https://osf.io/dmvst/?view_only=b50a8d3ec21b4a07b7977d0f56e79fc3. Additionally, all the analysis tables are provided as a supplement (S2 File).

## Results

### Study selection

We retrieved a total of 12, 986 bibliographic records from Scopus, PubMed, and Web of Science (Fig 1). Using Endnote and Rayyan, we identified and removed 7, 974 duplicates, resulting in 5, 012 unique records for title, abstract, and full text screening. We exported the unique records into Rayyan for the screening in two stages. Stage 1 involved a title and abstract screening in duplicate by the three reviewers and resulted 4, 211 excluded studies. The stage 2 screening, which involved screening the full text, led to 548 excluded studies.

**Fig 1.**
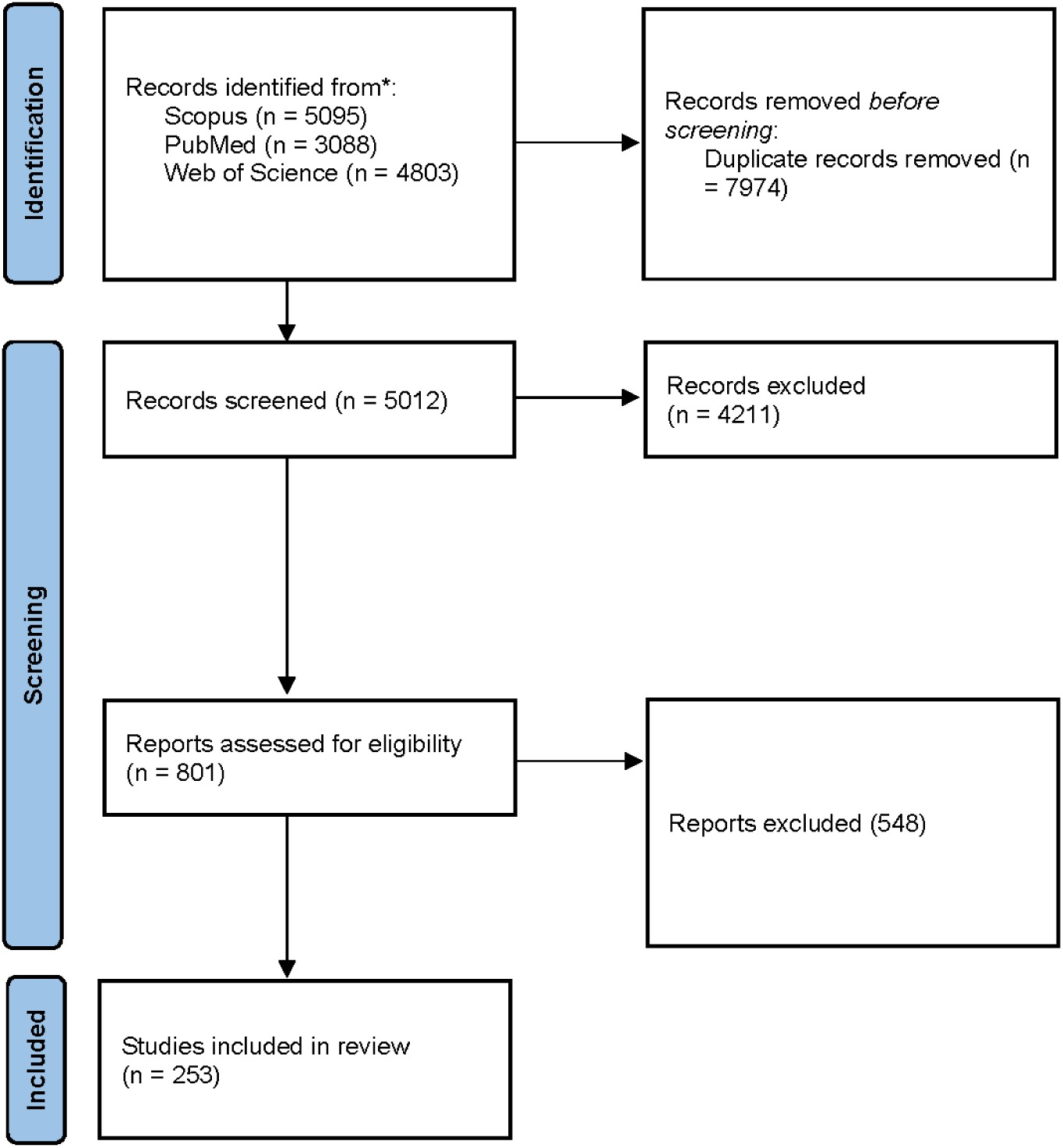
The PRISMA flow chart. Numbers described here include studies for both the human vaccine-preventable diseases and foot-and-mouth disease.

After the full text screening, 253 studies remained for the data extraction stage of which 228 on human vaccine-preventable diseases and 25 studies were on FMD. The FMD studies were only used for comparisons.

### Publications over time

Overall, a few outbreak response modelling papers were published between 1970-2005 (3.9%; 9/228), followed by a marked increase in publications until 2019 (96.1%; 220/228) (Fig 2).

**Fig 2.**
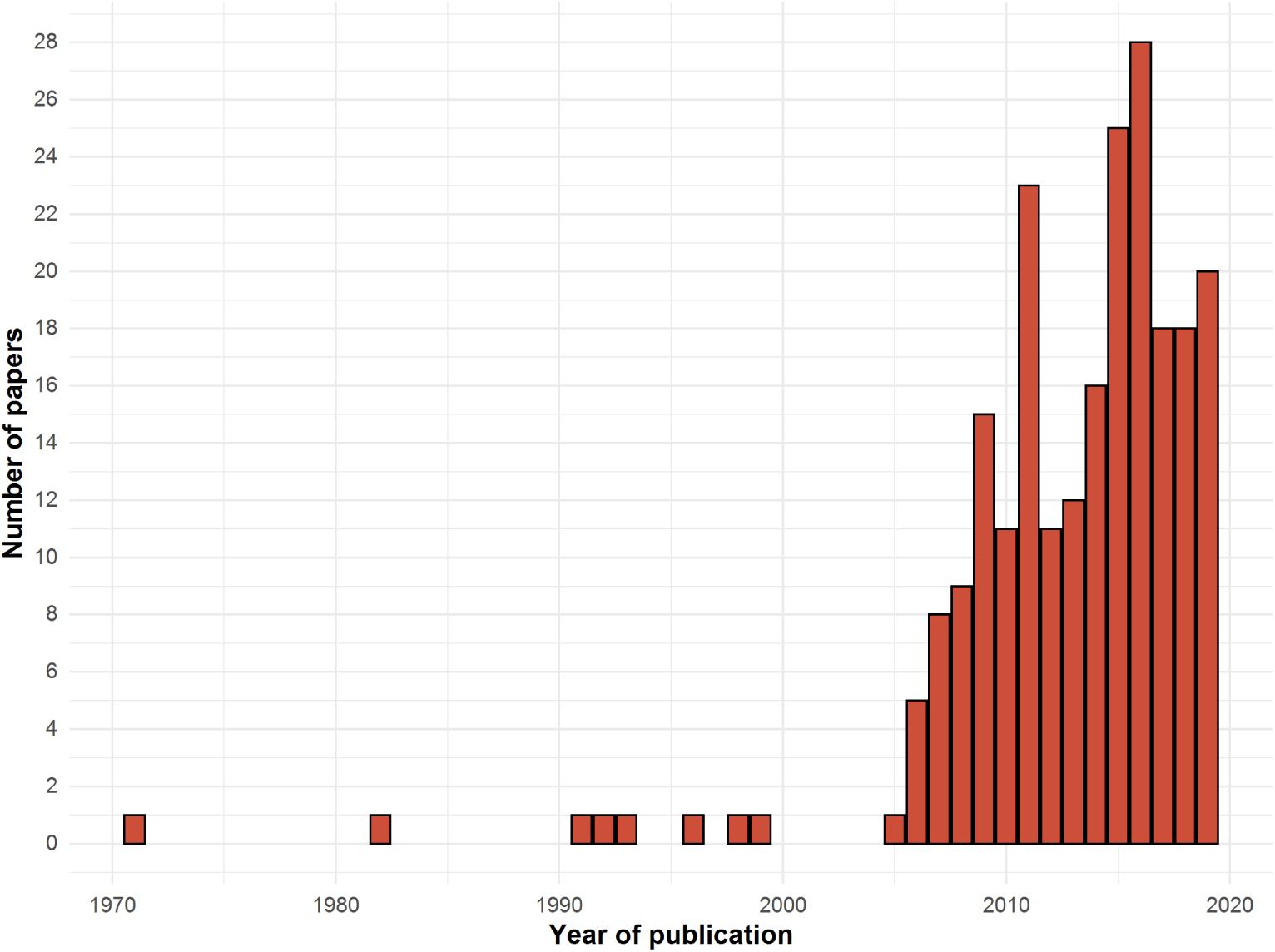
Number of publications per year (1970-2019).

We first categorised the papers according to the unique combinations of author affiliation types (Table 1).

Overall, papers with only authors from academic institution affiliations were the most common (56.1%; 128/228). Papers with author affiliations from academic and governmental organization affiliations were the second most common (25.0%; 57/228), and followed by those with academic and NGO affiliations (7.5%; 17/228). The least common were those with governmental and NGO affiliations (0.9%; 2/228).

As explained earlier, we ultimately grouped the author affiliation combinations into two collaboration types. There were more purely academic collaborations (56.1%; 128/228) than mixed collaborations (43.9%; 100/228).

To investigate the changes in collaboration types over time, we calculated the number and proportion of studies per collaboration type per year (Fig 3). In the past seven years (2013-2019), there was an absolute increase in the number of papers by both collaboration types. However, in the same period, there was no increase in the relative proportion of publications per year of mixed collaborations (S1 Fig).

**Fig 3.**
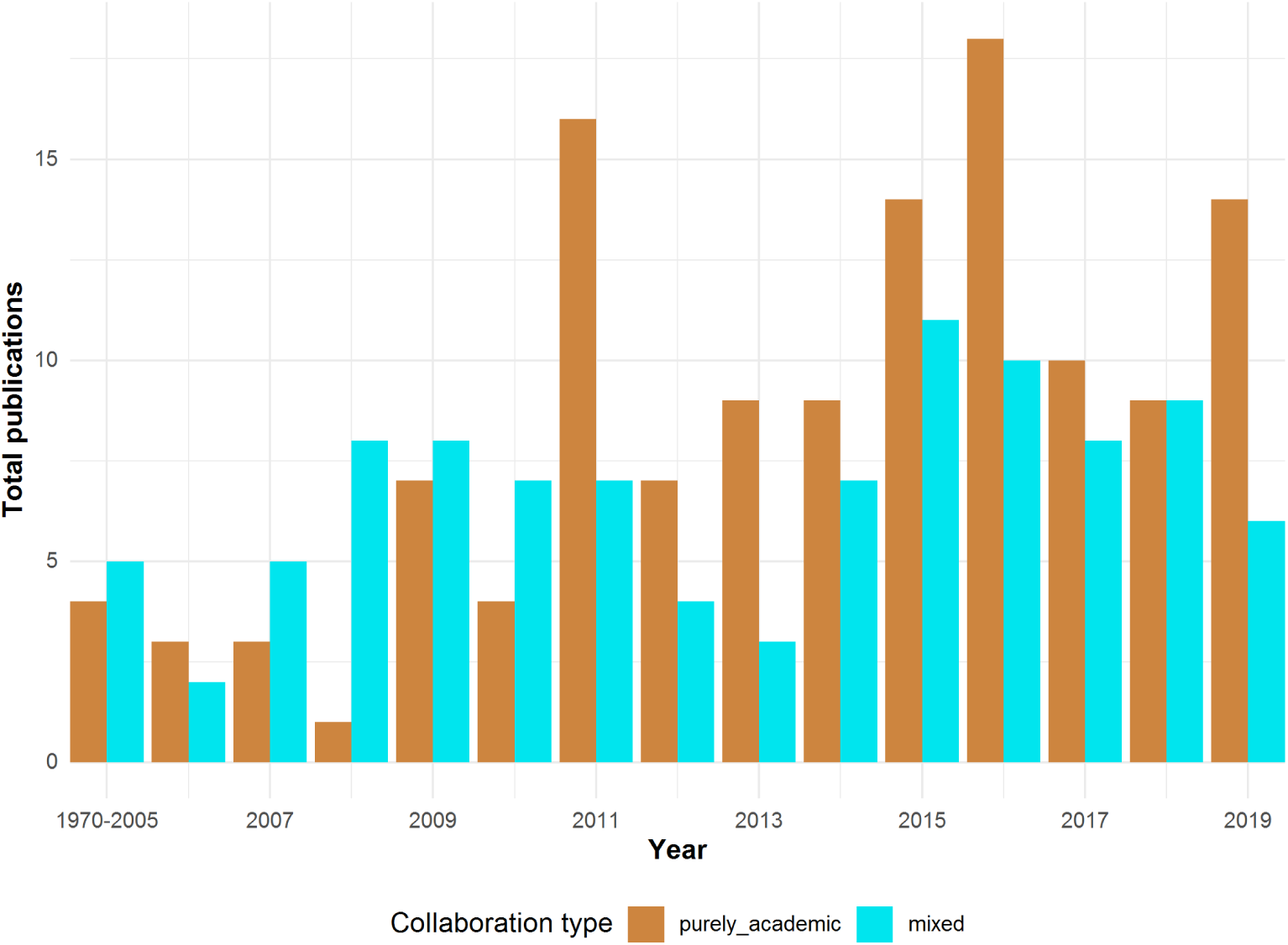
Total studies by collaboration type. The period from 1970-2005 has been lumped up due to the lack of publications. Academic collaborations refer to those authored by only authors with academic institution affiliations whereas mixed collaborations include those authored by a mixture of authors with academic institution affiliations, government institutions, and NGOs. Mixed collaborations are represented by the turquoise bars and purely academic collaborations by the brown bars.

### Locations studied

Overall, most of the papers were about actual locations (78.6%; 195/248). Among these, some studied a geographic location spanning more than one country but not classifiable as a continent (9.7%; 19/195). Among those 19 studies, West Africa was studied the most (47.4%; 9/19), followed by the whole globe (36.8%; 7/19), Southeast Asia (10.5%; 2/19), and the Northern Hemisphere (5.3%; 1/19).

When aggregated into continents, the Americas were studied the most (36.4%; 68/187), followed by Asia (25.1%; 47/187), Africa (24.6%; 46/187), Europe (13.4%; 25/187), and Oceania (0.5%; 1/187).

When disaggregated into countries, the United States was the most studied (21.1%; 37/176), followed by China (10.8%; 19/176), Canada (6.2%; 11/176), and Sierra Leone (5.7%; 10/176).

### Connection of authors to the location studied

We investigated the connection of the authors to the location studied and found that, overall, there were more studies with at least one author connected to the studied location (75.2%; 118/157) than not (24.8%; 39/157).

When stratified by collaboration types, mixed collaborations were more likely to have studies with at least one author in the location studied (83.1%; 63/77) compared with the purely academic collaborations (67.5%; 54/80).

### Diseases studied

Overall, the number of studies per disease was disproportionately distributed (Table 2). Influenza was the most studied (57.2%; 135/236), followed by Ebola (14.4%; 34/236), Dengue (5.1%; 12/236), and a tie between Cholera (4.7%; 11/236), and Measles (4.7%; 11/236).

**Table 2.** Number of studies per disease and collaboration type. Percentages are calculated from the row totals. The number of studies making up the percentages are shown in brackets. In counting the number of papers per disease studied, if a study was about more than one disease, we treated each instance as unique and counted them separately, leading to a total greater than the reported number of studies.

| Disease | Purely academic | Mixed | Total |
| --- | --- | --- | --- |
| Influenza | 52.6% (71) | 47.4% (64) | 135 |
| Ebola | 70.6% (24) | 29.4% (10) | 35 |
| Dengue | 50.0% (6) | 50.0% (6) | 12 |
| Cholera | 81.8% (9) | 18.2% (2) | 11 |
| Measles | 36.4% (4) | 63.6% (7) | 11 |
| Tuberculosis | 85.7% (6) | 14.3% (1) | 7 |
| Poliomyelitis | 28.6% (2) | 71.4% (5) | 7 |
| Varicella | 50.0% (2) | 50.0% (2) | 4 |
| Meningococcal meningitis | 33.3% (1) | 66.7% (2) | 3 |
| Pertussis | 50.0% (1) | 50.0% (1) | 2 |
| Pneumococcal disease | 50.0% (1) | 50.0% (1) | 2 |
| Yellow fever | 50.0% (1) | 50.0% (1) | 2 |
| Hepatitis A | 100.0% (1) | 0.0% (0) | 1 |
| Rubella | 100.0% (1) | 0.0% (0) | 1 |
| Typhoid | 100.0% (1) | 0.0% (0) | 1 |
| Hepatitis B | 0.0% (0) | 100.0% (1) | 1 |
| Malaria | 0.0% (0) | 100.0% (1) | 1 |
| Mumps | 0.0% (0) | 100.0% (1) | 1 |
| **Overall** | **55.5% (131)** | **44.5% (105)** | **100% (236)** |

When the diseases were broken down in terms of collaboration type, there were clear differences in their distribution. Influenza, Dengue, and Measles were more studied by mixed collaborations whereas Ebola and Cholera were more studied by purely academic collaborations.

### Intervention types and impact of vaccination

There were more papers about non-vaccination interventions (48.2%; 110/228), followed by those that modelled vaccination as part of a mix of interventions or do-nothing counterfactuals (41.7%; 95/228), and those that modelled vaccination as a single intervention for side-by-side comparison with other non-vaccination interventions (10.1%; 23/228).

The third group of papers that modelled vaccination as a single intervention allowed us to collate conclusions on the sole impact of vaccination. There were approximately the same number of studies in this set of studies belonging to the two collaborations types.

Concerning the conclusions about the sole impact of vaccination as a single intervention, most of the studies found vaccination to be the most impactful single intervention in side-by-side comparison with other non-vaccination interventions (82.6%; 19/23). Few studies found the vaccination to be less impactful compared to other interventions and for various reasons (17.4%; 4/23). Influenza was the disease studied in these latter four cases. Isolation, and antivirals were found to be more impactful than vaccination due to reasons including the delay until a strain-specific vaccine is developed to control the disease.

### Model structure, spatial heterogeneity, and model dynamics

There were more compartmental models (78.5%; 179/228) than agent-based models (20.2%; 46/228) with no clear difference in preference between the two collaboration types.

Approximately a third of the papers included spatial heterogeneity (28.9%; 66/228) with no clear difference between the two collaboration types.

Deterministic models were the most common (62.3%; 142/228) compared to stochastic (31.6%; 72/228) and hybrid models (6.1%; 14/228), that is models with both deterministic and stochastic components. Here, mixed collaborations were more likely to use stochastic models (40.0%; 40/100) than purely academic collaborations (25.0%; 32/128).

### Model parametrization and validation

The top three most commonly used parametrization methods included: combining literature sources and expert opinion/assumptions, literature sources and fitting to data, and literature sources only (Table 3).

**Table 3.** Model parametrization methods. We define parametrization as the method of determining the parameter values for the model. In the table, “Literature” means the model’s parameters were obtained from literature sources. “Expert opinion” means the values were assumed in consultation with experts of the field. “Fitted” means the model’s parameters were obtained through some form of mathematical or statistical fitting to a time series of data.

| Collaboration type | Literature and expert opinion | Literature and fitted | Literature | Expert opinion | Literature, expert opinion, and fitted | Fitted | Expert opinion and fitted | Total |
| --- | --- | --- | --- | --- | --- | --- | --- | --- |
| Academic | 27.3% (35) | 22.7% (29) | 18.0% (23) | 13.3% (17) | 9.4% (12) | 7.0% (9) | 2.3% (3) | 128 |
| Mixed | 25.0% (25) | 28.0% (28) | 15.0% (15) | 7.0% (7) | 19.0% (19) | 5.0% (5) | 1.0% (1) | 100 |
| Overall | 26.3% (60) | 25.0% (57) | 16.7% (38) | 10.5% (24) | 13.6% (31) | 6.1% (14) | 1.8% (4) | 228 |

The most common parametrization method among mixed collaborations was the combination of literature sources and fitting (27.3%; 35/128), whereas the combination of literature and expert opinion was the most common among purely academic collaborations (28.0%; 28/100).

Most studies did not perform any form of validation (63.2%; 144/228). Approximately a third used data to validate their models (34.6%; 79/228) (Table 4). Mixed collaborations were more likely to validate their model with data or the output of an independent model.

**Table 4.** Model validation methods. We define model validation as the method by which the model’s performance was measured. “None” means no validation was performed, or the model’s output was compared to another output from the same model. “Data” means the model was compared to independently observed data. “Another model’s output” means the model’s output was compared to an independent model’s output.

| Collaboration type | None | Data | Another model’s output | Data and another model | Total |
| --- | --- | --- | --- | --- | --- |
| Academic | 70.3% (90) | 28.9% (37) | 0.8% (1) | 0.0% (0) | 128 |
| Mixed | 54.0% (54) | 42.0% (42) | 3.0% (3) | 1.0% (1) | 100 |
| Overall | 63.2% (144) | 34.6% (79) | 1.8% (4) | 0.4% (1) | 228 |

### Data and model simulation code

More than half of the papers used datasets collected independent of the study for either the model parametrization or validation process (56.1%; 128/228). These papers were split approximately equally between the two collaboration types.

The use of accessible data was, however, different between the two groups. Purely academic collaborations were more likely to use data that could be accessed in the public domain (81.0%; 51/63) compared to mixed collaborations (56.9%; 37/65).

Few papers provided access to the model simulation code (1.7%; 4/228). Here, if a paper indicated that the authors could be contacted for the code, it was deemed as inaccessible due to the many hurdles with getting the code in time. Some papers reported the computer application/software/program/package used (R, Python, C++, Matlab, and so forth) but we did not collect that information.

### Patterns in the foot and mouth disease (FMD) literature

There were 25 studies about foot and mouth disease outbreaks. Due to the small number of studies, the absolute differences between the two collaboration types were generally not large enough to be considered (S2 File), hence, we report on the aggregated patterns.

In terms of collaboration types, slightly more than half of the 25 studies were authored by mixed collaborations (56.0%; 14/25) than purely academic collaborations (44.0%; 11/25). Almost all the papers had at least one author in the location studied (92.0%; 23/25).

Overall, vaccination was modelled as a single intervention for comparison in more than half of the studies (56.0%; 14/25), followed by vaccination in combination with other interventions (28.0%; 7/25), and no vaccination (16.0%; 4/25). Few of the first set of studies found vaccination to be the most impactful as a single intervention (14.3%; 2/14).

In general, the FMD models had more agent-based (76.0%; 19/25) and spatially explicit structure (72.0%; 18/25), largely used stochastic dynamics (56.0%; 14/25), and performed more sensitivity analyses (56.0%; 14/25).

In terms of model parametrization, the most common method was the combined use of literature sources and expert opinion (28.0%; 7/25), followed by fitting to data only (20.0%; 5/25), and the combined use of literature sources, expert opinion, and fitting to data (16.0%; 4/25). Here, mixed collaborations were more likely than purely academic collaborations to combine literature sources and expert opinion (42.9%6/14). Furthermore, most of the FMD models were not validated (72.0%; 18/25).

## Discussion

For the period 1970 – 2019, co-authorship by purely academic collaborations formed over 56% of the outbreak response impact modelling studies of human vaccine-preventable diseases. Both collaboration types increased in absolute numbers over the past seven years. Regarding modelling practices between the two collaboration types, mixed collaborations were more likely to: (i) include authors affiliated through their institutions to the location studied, (ii) use more complex modelling practices including stochastic model dynamics, parametrization methods involving a combination of literature sources, expert opinion/assumptions, and fitting to data, and validate their models with data collected independent of the study. Even though this review was about human vaccine-preventable diseases, only 51.8% of the studies modelled vaccination in some form. Moreover, only 10.1% of the total studies modelled vaccination for comparison with other non-vaccination strategies in the same analysis. Influenza was disproportionately the most studied disease, followed by Ebola, and Dengue.

There were several differences in models and practices between the human disease literature and foot and mouth disease (FMD). Mixed collaborations dominated the FMD literature, with almost all papers having an author in the location studied. Furthermore, most of the FMD models and modelling practices were more complex involving the use of spatial, stochastic, agent-based models. However, the FMD models were less likely to be validated with real world data.

Collaborations, especially between authors from academic institutions, governmental, and non-governmental organizations responsible for outbreak response, could lead to knowledge transfer and improved decision-making [16]. Moreover, such diversity in modelling collaborations could expand the skill sets needed to solve the complex real world outbreak response needs. However, it is well known that there is a weak link between academic research and decision-making in general, and research findings are often not easily translated into decisions [16–18]. This gap in knowledge translation exists especially between outbreak response researchers and public health decision-makers [16, 19]. There are examples of the commissioning of modelling by organisations such as the US Centers for Disease Control and Prevention (CDC) and the World Health Organization (WHO) to inform decision-making during past outbreaks of Influenza, Ebola, Zika, and Dengue [16]. However, many decision-makers remain cautious about the uptake of modelling due, in part, to issues about transparency in assumptions, credibility and ease of use of modelling software, and adaptability of the results to other settings and existing policies [16].

To bridge this knowledge translation gap between modellers and decision-makers, modelling collaborations that deepen the interaction between model developers and decision-making have been proposed [17, 19]. This could be achieved through the sharing of expertise so that the academics formulate, parametrize, and validate the models taking into account the expert opinions of the decision-makers so that the findings are more tailored towards implementation. Mixed or interdisciplinary collaborations have been reported to be strongly associated with research impact and translation to decision-making [18]. Hence, outbreak response, which is broadly an operational field, would ideally have modelling groups/collaborations that are comprised of academics and decision-makers to bridge this gap. In this review, we observed an absolute increase in the number of papers published by mixed collaborations (Fig 3). This could suggest an increase in the uptake or recognition of modelling as an outbreak response decision-making tool. Future research to investigate and explain this increase could advance our understanding of the contribution of modelling to outbreak response decision-making.

The inclusion of local experts in outbreak response modelling teams helps with more tailored problem-solving and decision-making and better reception of the decisions made [7]. In that regard, mathematical modelling collaborations involving local experts or, in this review, locally affiliated co-authors are an important first step towards achieving this. We found that most of the human disease studies had authors with an institutional affiliation in the location studied. We observed the same pattern for FMD albeit at a much higher percentage. However, when the human disease data were disaggregated, the results showed that mixed collaborations had a much higher percentage of studies with at least one author in the studied location compared to purely academic collaborations. Coupling this with the absolute increase in publications by mixed collaborations in the past seven years could suggest an increase in the uptake of modelling as a public health decision-making tool. However, we did not measure this causality. Research that measures the direct impact of mathematical modelling in public health decision-making remains lacking in the literature [16]. Future studies designed to investigate this uptake of models and whether it has had an impact on outbreak response would contribute meaningfully to the field of outbreak response modelling.

Three classes of interventions with respect to vaccination were modelled: non-vaccination (mostly antiviral use and non-pharmaceutical interventions), vaccination mixed with other complementary non-vaccination interventions, and vaccination as a single intervention for side-by-side comparison with other non-vaccination alternatives. The last class of interventions were the only eligible set for our analysis of the conclusions about whether vaccination is always the most impactful intervention when modelling is used as the assessment tool. Even though all the reviewed studies were about human vaccine-preventable diseases, only 10.1% of the studies modelled vaccination as a single intervention for side-by-side comparison with non-vaccination interventions. This lack of studies assessing the sole impact of vaccination reflects the reality that vaccination is often implemented alongside other interventions and never alone, making it difficult to assess its sole impact. Nevertheless, mathematical modelling is an ethically viable way to assess the sole impact of vaccination, and hence, an essential tool for decision-making.

Among the studies that modelled vaccination as a single intervention for comparison with other single non-vaccination interventions, vaccination was found to be the most impactful intervention in 82% of the studies. Four studies out of the 23 that modelled influenza found case isolation [20–22] and antiviral prophylaxis and/or therapeutics [23] to be more impactful than vaccination assuming those interventions could be implemented immediately, given significantly efficient isolation and large antiviral stockpiles before the outbreak/pandemic. Among these four studies, some common assumptions that influenced how vaccination was implemented included: the late arrival of effective vaccines [20], low versus high vaccination rates [21], the use of partially effective pre-existing vaccines against the current strain reactively [24] or pre-emptively [25]. In all four studies, it was, however, recommended that, vaccination be used in combination with the alternative interventions. Even though antivirals were found to be more impactful in some cases, its prolonged use was discouraged to prevent the development of antiviral resistance [24]. On the other hand, there was a high percentage of studies in the FMD literature about the use of vaccination as a single intervention and vaccination was rarely found to be the most impactful intervention.

We found some commonalities and differences in the choice of model structure and dynamics between the two collaboration types in the human disease literature. Mixed collaborations were more likely to use “complex” models and practices. We are not making any value judgements with regards to complexity but are only highlighting these differences that might require further investigation. Moreover, it is common to model an outbreak using alternative model choices and assumptions depending on the question being answered [26]. However, the results must always be interpreted with cognisance of the limitations/assumptions of the model. The use of more complex models and practices by mixed collaborations is likely due to the fact that often, real world public health policy-related decision-making is operational, requiring finer details in models and approaches to answer the questions posed in a practical way. Our definition of mixed collaborations meant that it may have involved decision-makers. It is, therefore, likely that their need for practical solutions could have influenced higher levels of model details or complexity.

The FMD models were generally more complex than the human disease models. This is not surprising because the nature of FMD spread often requires the inclusion of farm structure, farm connectivity, and demographics to capture the disease’s dynamics accurately [27]. Future studies designed to explain the differences in modelling practices of human outbreak response modelling groups might help to explain what we have observed in this review.

A little over half of the human disease papers used observed data for parametrization or validation. Additionally, only 4 studies shared their code in an easy to access form. On the other hand, the FMD models were generally not validated with observed data. Owing to this, it might be difficult to reproduce some of the results in these outbreak response modelling studies. We recognise that data sharing in public health raises a lot of debate regarding privacy and intellectual property, and often, authors are hindered by institutional data sharing policies [28]. We, however, recommend that data and code be shared where possible to promote open science practices that help advance the field.

This review had several limitations. First, outbreak response models and analyses are not always published in peer-reviewed journals, but this systematic review only focussed on peer-reviewed articles. It is, therefore, possible that some relevant studies were missed by our search strategy. Second, we used the author list and affiliations to classify studies as either purely academic or mixed. However, this could cause some mixed collaborations to be misclassified as purely academic, especially in cases where non-academics contributed to papers classified as purely academic but were not included on the author list. It is, however, standard practice to include individuals who contributed substantially to a piece of scientific writing, hence, if that was not done for a paper, it could imply that the level of interaction was not high enough to warrant the credit of authorship. Third, in the absence of an explicit measure of contribution of mathematical modelling to outbreak response decision-making, we used mixed collaborations as a proxy. Thus, we may be under-estimating the number of mixed collaborations in the literature. Forth, we only surveyed the literature on a specific study design – mechanistic models. We excluded statistical models and other computational models, which are not mechanistic. Hence, the results of this systematic review should only be interpreted in the context of the mechanistic modelling landscape. Furthermore, it is also possible that some purely academic collaborations contribute to decision-making whereas some mixed collaborations are purely an academic exercise. Also, we conducted our database searches in January 2020 and therefore do not reflect the literature, or any changes in practice, associated with the COVID-19 pandemic. Lastly, by only including papers published in English, we most certainly missed papers published in other languages.

Numerous factors could explain the patterns we have observed in this review, and we would recommend future studies that will use appropriate study designs, for example, interviews of modelling groups and public health decision-makers, to explain why certain model choices and modelling practices were made by the collaboration types. Future studies should investigate when modelling results directly impacted decision-making and what determined that to identify best practices that will strengthen the link between modelling and decision-making in the future. We expect that collaboration between academia, decision-makers, and local experts will enhance decision-making by accounting for aspects of policy and decision-making that might be overlooked in studies conducted mainly as a theoretical exercise.

## Data Availability

All data extracted from the studies are available from at the following link https://osf.io/dmvst/?view_only=b50a8d3ec21b4a07b7977d0f56e79fc3

## Other information

A protocol following the PRISMA guidelines for systematic review protocols and outlining the procedures for this systematic review was registered on PROSPERO with registration number CRD42020160803 and published through a peer-reviewed process [29].

## Supporting information

**S1 File Search strings and results, data items extracted, and questionnaire for data extraction**.

**S1 Fig Collaboration patterns in time (proportions per year)**.

**S2 File Analysis of extracted data**.

**S3 Table PRISMA checklist**.

## References

1. Sigfrid L, Maskell K, Bannister PG, Ismail SA, Collinson S, Regmi S, et al. Addressing challenges for clinical research responses to emerging epidemics and pandemics: a scoping review. BMC medicine. 2020;18(1):1–15.

2. Kretzschmar M. Disease modeling for public health: added value, challenges, and institutional constraints. Journal of Public Health Policy. 2020;41(1):39–51. doi:10.1057/s41271-019-00206-0s.

3. van Kerkhove MD, Ferguson NM. Epidemic and intervention modeling - a scientific rationale for policy decisions? Lessons from the 2009 influenza pandemic. Bulletin of the World Health Organization. 2012;90(4):306–310. doi:10.2471/BLT.11.097949.

4. Okiror S, Toure B, Davis B, Hydarov R, Ram B, Nwogu C, et al. Lessons Learnt from Interregional and Interagency Collaboration in Polio Outbreak Response in the Horn of Africa. Journal of Immunological Sciences. 2021;Special Is(2):40–47. doi:10.29245/2578-3009/2021/s2.1112.

5. Whitty CJM, Farrar J, Ferguson N, Edmunds WJ, Piot P, Leach M, et al. Infectious disease: Tough choices to reduce Ebola transmission. Nature. 2014;515(7526):192–194. doi:10.1038/515192a.

6. Heesterbeek H, Anderson RM, Andreasen V, Bansal S, DeAngelis D, Dye C, et al. Modeling infectious disease dynamics in the complex landscape of global health. Science. 2015;347(6227). doi:10.1126/science.aaa4339.

7. Abramowitz SA, Bardosh KL, Leach M, Hewlett B, Nichter M, Nguyen VK. Social science intelligence in the global Ebola response. The Lancet. 2015;385(9965):330. doi:10.1016/S0140-6736(15)60119-2.

8. Lofgren E, Halloran ME, Rivers CM, Drake JM, Porco TC, Lewis B, et al. Opinion: Mathematical models: A key tool for outbreak response. Proceedings of the National Academy of Sciences. 2015;112(2):E234–E234. doi:10.1073/pnas.1423846112.

9. Abramo G, D’Angelo CA, Di Costa F. Do interdisciplinary research teams deliver higher gains to science? Scientometrics. 2017;111(1):317–336. doi:10.1007/s11192-017-2253-x.

10. Wagner CS, Roessner JD, Bobb K, Klein JT, Boyack KW, Keyton J, et al. Approaches to understanding and measuring interdisciplinary scientific research (IDR): A review of the literature. Journal of informetrics. 2011;5(1):14–26.

11. Littmann M, Selig K, Cohen-Lavi L, Frank Y, Hönigschmid P, Kataka E, et al. Validity of machine learning in biology and medicine increased through collaborations across fields of expertise. Nature Machine Intelligence. 2020;doi:10.1038/s42256-019-0139-8.

12. Page MJ, McKenzie JE, Bossuyt PM, Boutron I, Hoffmann TC, Mulrow CD, et al. The PRISMA 2020 statement: an updated guideline for reporting systematic reviews. International Journal of Surgery. 2021;88:105906.

13. Lessler J, Cummings DAT. Mechanistic models of infectious disease and their impact on public health. American Journal of Epidemiology. 2016;183(5):415–422. doi:10.1093/aje/kww021.

14. Reiner RC, Perkins TA, Barker CM, Niu T, Chaves LF, Ellis AM, et al. A systematic review of mathematical models of mosquito-borne pathogen transmission: 1970-2010. Journal of The Royal Society Interface. 2013;10(81):20120921–20120921. doi:10.1098/rsif.2012.0921.

15. R Core Team. R: A Language and Environment for Statistical Computing; 2021. Available from: https://www.R-project.org/.

16. Muscatello DJ, Chughtai AA, Heywood A, Gardner LM, Heslop DJ, MacIntyre CR. Translation of real-time infectious disease modeling into routine public health practice. Emerging infectious diseases. 2017;23(5).

17. Choi BC, Pang T, Lin V, Puska P, Sherman G, Goddard M, et al. Can scientists and policy makers work together? Journal of Epidemiology & Community Health. 2005;59(8):632–637.

18. Deelstra Y, Nooteboom SG, Kohlmann HR, Van Den Berg J, Innanen S. Using knowledge for decision-making purposes in the context of large projects in The Netherlands. Environmental Impact Assessment Review. 2003;23(5):517–541. doi:10.1016/S0195-9255(03)00070-2.

19. Rivers C, Chretien JP, Riley S, Pavlin JA, Woodward A, Brett-Major D, et al. Using “outbreak science” to strengthen the use of models during epidemics. Nature communications. 2019;10(1):1–3.

20. Yasuda H, Suzuki K. Measures against transmission of pandemic H1N1 influenza in Japan in 2009: simulation model. Eurosurveillance. 2009;14(44):19385.

21. Gao X, Wei J, Cowling BJ, Li Y. Potential impact of a ventilation intervention for influenza in the context of a dense indoor contact network in Hong Kong. Science of the Total Environment. 2016;569-570:373–381. doi:10.1016/j.scitotenv.2016.06.179.

22. Chen T, Zhao B, Liu R, Zhang X, Xie Z, Chen S. Simulation of key interventions for seasonal influenza outbreak control at school in Changsha, China. Journal of International Medical Research. 2020;48(1):0300060518764268.

23. Gumel AB, Nuño M, Chowell G. Mathematical assessment of Canada’s pandemic influenza preparedness plan. Canadian Journal of Infectious Diseases and Medical Microbiology. 2008;19(2):185–192.

24. Gumel AB. Global dynamics of a two-strain avian influenza model. International Journal of Computer Mathematics. 2009;86(1):85–108. doi:10.1080/00207160701769625.

25. Chen CJ, Lin TY, Huang YC. Letter to the editor: Occurrence of modified measles during outbreak in Taiwan in 2018. Eurosurveillance. 2018;23(37):1–2. doi:10.2807/1560-7917.ES.2018.23.37.1800485.

26. Basu S, Andrews J. Complexity in mathematical models of public health policies: a guide for consumers of models. PLoS medicine. 2013;10(10):e1001540.

27. Kinsley A, VanderWaal K, Craft M, Morrison R, Perez A. Managing complexity: Simplifying assumptions of foot-and-mouth disease models for swine. Transboundary and emerging diseases. 2018;65(5):1307–1317.

28. Kim Y, Stanton JM. Institutional and individual factors affecting scientists’ data-sharing behaviors: A multilevel analysis. Journal of the Association for Information Science and Technology. 2016;67(4):776–799.

29. Azam JM, Are EB, Pang X, Ferrari MJ, Pulliam JRC. Outbreak response intervention models of vaccine-preventable diseases in humans and foot-and-mouth disease in livestock: a protocol for a systematic review. BMJ open. 2020;10(10):e036172. doi:10.1136/bmjopen-2019-036172.

