## Supplementary figures and images for "Models and modelling practices for assessing the impact of outbreak response interventions to human vaccine-preventable diseases (1970-2019) - A systematic review"

### S1 Fig

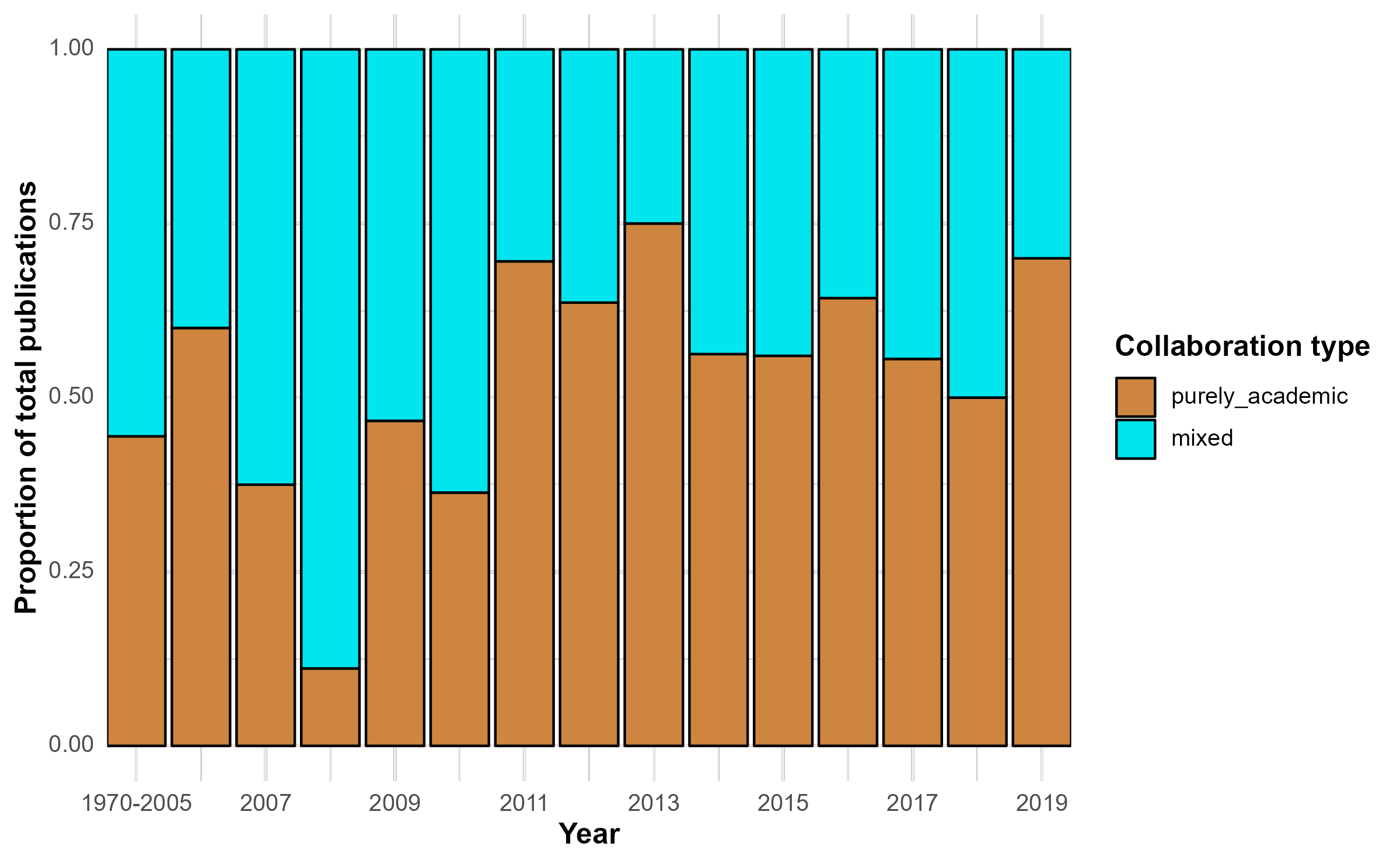
