## Supplementary material for "Models and modelling practices for assessing the impact of outbreak response interventions to human vaccine-preventable diseases (1970-2019) - A systematic review": S1 File

March 31, 2022

#### 1 Human vaccine-preventable diseases considered

Table 1: The World Health Organization (WHO) list of diseases with an available vaccine as of Jan 1, 2020.

| | |
|---|---|
| Cholera | Mumps |
| Dengue | Pertussis |
| Diphtheria | Pneumococcal disease |
| Hepatitis A | Poliomyelitis |
| Hepatitis B | Rabies |
| Hepatitis E | Rotavirus |
| Haemophilus influenzae type b (Hib) | Rubella |
| Human papillomavirus (HPV) | Tetanus |
| Influenza | Tick-borne encephalitis |
| Japanese encephalitis | Tuberculosis |
| Malaria | Typhoid |
| Measles | Varicella |
| Meningococcal meningitis | Yellow Fever |

#### 2 Data items

The following information will be extracted from each of the included studies:

1. Disease(s) modelled
2. Study's main objective(s)
3. Location studied and whether the first or last author has an affiliation in the location
4. Model dynamics: deterministic, stochastic, or a hybrid
5. How individuals are represented in model: individual-bases or compartmental
6. Whether spatial structure was included
7. Interventions modelled: vaccination (ring, mass, targeted, prophylactic, pulse) or non-vaccination (movement restrictions, palliative care, quarantine, isolation, treatment, other(s) as indicated)

8. Model parametrisation (We define model parametrization as the process of obtaining the model's parameter values either from literature or through some kind of mathematical or statistical technique)
9. Method of model validation with some form of data. (We define model validation as any procedure used to evaluate the model's suitability for the data and problem being studied)
10. Outcome measured with model
11. Authors' conclusion on the predicted or potential impact of the intervention
12. Timing of modelling practice: retrospective, real-time, other(s) as indicated
13. Sensitivity analysis performed or not
14. Data and code availability

### 3 Questionnaire for data extraction

#### 3.1 Citation information

1. What is the title of this paper?
2. What is the year of publication?
3. What is the DOI of the paper?

#### 3.2 Collaboration

4. Which of the following describe the nature of the author affiliations?
    - Academic institutions (universities, research institutions, etc)
    - Government institutions (government ministries or departments, etc)
    - Non-governmental institutions
5. Which country is this study based on? [Dropdown menu of all countries including an option for hypothetical, and other, if multiple. Please specify]
6. Are any of the author affiliations situated in the country being studied?
    - Yes
    - No

#### 3.3 Introduction

##### 3.3.1 Disease

7. Which disease(s) was modelled in this study?

8. Which of the following will describe the study's broad objective(s)?

    - Assessing the potential impact/effectiveness of past interventions
    - Assessing the potential impact/effectiveness of future/proposed interventions
    - Other. Please indicate.

#### 3.4 Methods

##### 3.4.1 Model structure

9. How are individuals represented in the model?

    - Compartments, where identical individuals are grouped according to disease state
    - Agent/Individual-based, where each agent is captured individually

10. Is space explicitly represented in the model?

    - Yes
    - No
    - Unclear. Please indicate why.

11. Which of the following describes the model structure?

    - Deterministic
    - Stochastic
    - Other. Please indicate

##### 3.4.2 Calibration and validation

12. How were the model's parameter values determined?

    - Values were sourced from various literature sources, including, systematic reviews studies reporting on primary data
    - Values were assumed/guessed based on a plausible range (expert opinion)
    - Values were fitted to a time series

13. Which of the following best describe the model validation process, that is, the process of evaluating the model's ability to describe the dynamics of the disease?

    - The model was not validated. (This includes where one set of the model's runs were compared to another set of runs from the same model.)

- The model's output was compared to an independently observed dataset
- The model's output was compared to at least one other model's output
- Other. Please indicate

###### 3.4.3 Outbreak and intervention

14. What type of outbreak was this study based on?
    - A real outbreak that occurred
    - A hypothetical outbreak
15. Which of the following would describe the type of intervention modelled?
    - Antiviral prophylaxis
    - Bednets
    - Behaviour change
    - Contact tracing
    - Culling
    - Education
    - Face masks
    - Handwashing
    - Hospital bed capacity
    - Improved hygiene
    - Indoor/outdoor spraying
    - Isolation
    - Larvicides
    - Media campaign
    - Palliative care
    - Protective clothing (PPE, other than face masks)
    - Quarantine
    - Safe burial
    - Social/physical distancing
    - School closure
    - Testing/screening
    - Travel restriction/border control
    - Treatment
    - Travel screening
    - Vaccination
    - Ventilation
    - Workplace closure
    - Other. Please indicate.

###### 3.4.4 Impact of interventions

16. Was vaccination modelled for comparison with other interventions?
    - Yes
    - No
17. If yes, was vaccination found to be the most effective strategy in terms of the stated objective?
    - Yes
    - No
    - The outcomes were inconclusive
18. What was the timing of the modelling practice?

- Prospective
- Retrospective
- In real-time during the outbreak

19. Which outcome was measured?

- Cases averted
- Intervention coverage
- Lives saved
- Deaths prevented/averted
- Campaign duration
- Outbreak duration/timing
- Final epidemic size/Outbreak size/Total cases
- Cost (direct cost/cost-effectiveness/cost-benefit)
- Timing of peak
- Attack rate
- Hospitalizations
- QUALYS
- DALYs
- Case-fatality
- Other. Please indicate

###### 3.4.5 Sensitivity analysis

20. Was a sensitivity analysis performed?

- Yes
- No

###### 3.4.6 Reproducibility/transparency

**Data**

21. Was any independently observed data used in this modelling study? (Independently observed data implies data that was not simulated with the model being used for evaluation in this study.)

- No
- Yes

22. If Yes, Is the independently observed data openly available?

- Yes
- No
- Nothing about this was mentioned

###### 3.4.7 Code

23. Is the simulation code openly available?

- Yes (supplementary materials, git, zenodo, google drive, etc)
- No, including studies in where the authors indicated they could be emailed for the simulation code

### 4 Search strings and results

Below are the detailed search strategies and results for the three bibliographic databases used:

Table 2: Scopus search strings and results.

| Search | Query | Records found |
|---|---|---|
| #1 | TITLE-ABS-KEY ( epidemic OR outbreak OR emergency OR reactive OR crisis ) | 1,834,598 |
| #2 | TITLE-ABS-KEY ( respon* OR manage* OR control OR interven* OR strateg* ) | 19,366,062 |
| #3 | TITLE-ABS-KEY ( stochastic OR transmission OR computational OR mathematical OR mechanistic OR statistical OR simulation OR "In silico" OR dynamic* ) | 11 890 240 |
| #4 | TITLE-ABS-KEY ( model* ) | 12 504 649 |
| #5 | TITLE-ABS-KEY ( cholera OR dengue OR diphtheria OR ebola OR "Foot-and-mouth" OR "foot and mouth" OR fmd OR "Hepatitis A" OR "Hepatitis B" OR "Hepatitis E" OR "Haemophilus influenzae type b" OR hib OR "Human papillomavirus" OR hpv OR influenza ) ) OR ( TITLE-ABS-KEY ( "Japanese encephalitis" OR malaria OR measles OR "Meningococcal meningitis" OR mumps OR pertussis OR "Whooping cough" OR "Pneumococcal disease" OR poliomyelitis OR polio OR rabies OR rotavirus OR rubella ) ) OR ( TITLE-ABS-KEY ( tetanus OR "Tick-borne encephalitis" OR tuberculosis OR typhoid OR varicella OR chickenpox OR "Yellow Fever" OR "vaccine-preventable" ) ) | 1 123 110 |
| #6 | TITLE-ABS-KEY ( epidemic OR outbreak OR emergency OR reactive OR crisis ) ) AND ( TITLE-ABS-KEY ( respon* OR manage* OR control OR interven* OR strateg* ) ) AND ( TITLE-ABS-KEY ( stochastic OR transmission OR computational OR mathematical OR mechanistic OR statistical OR simulation OR "In silico" OR dynamic* ) ) AND ( TITLE-ABS-KEY ( model* ) ) AND ( ( TITLE-ABS-KEY ( cholera OR dengue OR diphtheria OR ebola OR "Foot-and-mouth" OR "foot and mouth" OR fmd OR "Hepatitis A" OR "Hepatitis B" OR "Hepatitis E" OR "Haemophilus influenzae type b" OR hib OR "Human papillomavirus" OR hpv OR influenza ) ) OR ( TITLE-ABS-KEY ( "Japanese encephalitis" OR malaria OR measles OR "Meningococcal meningitis" OR mumps OR pertussis OR "Whooping cough" OR "Pneumococcal disease" OR poliomyelitis OR polio OR rabies OR rotavirus OR rubella ) ) OR ( TITLE-ABS-KEY ( tetanus OR "Tick-borne encephalitis" OR tuberculosis OR typhoid OR varicella OR chickenpox OR "Yellow Fever" OR "vaccine-preventable" ) ) ) | 5 253 (5 095*) |

*Scopus returned 5 253 records but we could not retrieve 158 records after several attempts.

Table 3: PubMed search strings and results.

| Search | Query | Records found |
|---|---|---|
| #1 | Search (Epidemic OR Outbreak OR Emergency OR Reactive OR Crisis) | 1 021 994 |
| #2 | Search (respon* OR manage* OR control OR interven* OR strateg*) | 9 134 363 |
| #3 | Search (Stochastic OR Transmission OR Computational OR Mathematical OR Mechanistic OR Statistical OR Simulation OR "In silico" OR Dynamic*) | 2 563 238 |
| #4 | Search (model*) | 2 222 906 |
| #5 | Search (Cholera OR Dengue OR Diphtheria OR Ebola OR "Foot-and-mouth" OR "foot and mouth" OR FMD OR "Hepatitis A" OR "Hepatitis B" OR "Hepatitis E" OR "Haemophilus influenzae type b" OR Hib OR "Human papillomavirus" OR HPV OR Influenza OR "Japanese encephalitis" OR Malaria OR Measles OR "Meningococcal meningitis" OR Mumps OR Pertussis OR "Whooping cough" OR "Pneumococcal disease" OR Poliomyelitis OR Polio OR Rabies OR Rotavirus OR Rubella OR Tetanus OR "Tick-borne encephalitis" OR Tuberculosis OR Typhoid OR Varicella OR Chickenpox OR "Yellow Fever" OR "vaccine-preventable") | 887 440 |
| #6 | Search ((((((Epidemic OR Outbreak OR Emergency OR Reactive OR Crisis))) AND ((Response OR Management OR Control OR Intervention OR Strategies))) AND ((Stochastic OR Transmission OR Computational OR Mathematical OR Mechanistic OR Statistical OR Simulation OR "In silico" OR Dynamic*))) AND model*) AND ((Cholera OR Dengue OR Diphtheria OR Ebola OR "Foot-and-mouth" OR "foot and mouth" OR FMD OR "Hepatitis A" OR "Hepatitis B" OR "Hepatitis E" OR "Haemophilus influenzae type b" OR Hib OR "Human papillomavirus" OR HPV OR Influenza OR "Japanese encephalitis" OR Malaria OR Measles OR "Meningococcal meningitis" OR Mumps OR Pertussis OR "Whooping cough" OR "Pneumococcal disease" OR Poliomyelitis OR Polio OR Rabies OR Rotavirus OR Rubella OR Tetanus OR "Tick-borne encephalitis" OR Tuberculosis OR Typhoid OR Varicella OR Chickenpox OR "Yellow Fever" OR "vaccine-preventable")) | 3 088 |

Table 4: Web of Science search strings and results

| Search | Query | Records found |
|---|---|---|
| # 1 | TOPIC: (Epidemic OR Outbreak OR Emergency OR Reactive OR Crisis) | 1 241 675 |
| # 2 | TOPIC: (respon* OR manage* OR control OR interven* OR strateg*) | 13 791 565 |
| # 3 | TOPIC: (Stochastic OR Transmission OR Computational OR Mathematical OR Mechanistic OR Statistical OR Simulat* OR "In silico" OR Dynamic*) | 7 869 289 |
| # 4 | TOPIC: (model*) | 705 715 |
| # 5 | TOPIC: (Cholera OR Dengue OR Diphtheria OR Ebola OR "Foot-and-mouth" OR "foot and mouth" OR FMD OR "Hepatitis A" OR "Hepatitis B" OR "Hepatitis E" OR "Haemophilus influenzae type b" OR Hib OR "Human papillomavirus" OR HPV OR Influenza OR "Japanese encephalitis" OR Malaria OR Measles OR "Meningococcal meningitis" OR Mumps OR Pertussis OR "Whooping cough" OR "Pneumococcal disease" OR Poliomyelitis OR Polio OR Rabies OR Rotavirus OR Rubella OR Tetanus OR "Tick-borne encephalitis" OR Tuberculosis OR Typhoid OR Varicella OR Chickenpox OR "Yellow Fever" OR "vaccine-preventable") | 8 790 133 |
| #6 | TOPIC: (Epidemic OR Outbreak OR Emergency OR Reactive OR Crisis) AND TOPIC: (Respon* OR Manage* OR Control OR Interven* OR Strateg*)) AND TOPIC: (Stochastic OR Transmission OR Computational OR Mathematical OR Mechanistic OR Statistical OR Simulation OR In silico OR Dynamic*) AND TOPIC: (model*) AND TOPIC: (Cholera OR Dengue OR Diphtheria OR Ebola OR "Foot-and-mouth" OR "foot and mouth" OR FMD OR "Hepatitis A" OR "Hepatitis B" OR "Hepatitis E" OR "Haemophilus influenzae type b" OR Hib OR "Human papillomavirus" OR HPV OR Influenza OR "Japanese encephalitis" OR Malaria OR Measles OR "Meningococcal meningitis" OR Mumps OR Pertussis OR "Whooping cough" OR "Pneumococcal disease" OR Poliomyelitis OR Polio OR Rabies OR Rotavirus OR Rubella OR Tetanus OR "Tick-borne encephalitis" OR Tuberculosis OR Typhoid OR Varicella OR Chickenpox OR "Yellow Fever" OR "vaccine-preventable") | 4 803 |
