## Supplementary material for "Models and modelling practices for assessing the impact of outbreak response interventions to human vaccine-preventable diseases (1970-2019) - A systematic review": S2 File

### Analysis of the included studies

#### Contents

**Human vaccine-preventable diseases** 1
Collaboration types (aggregated) 1
Locations studied 2
Geographic connection of authors to the studied locations 4
Interventions 5
Modelling objectives 6
Outbreak types 6
Individual heterogeneity: agent-based versus compartmental models 6
Spatial heterogeneity 6
Model dynamics: deterministic vs stochastic 7
Outcomes measured 7
Sensitivity analysis 9
Data use and data availability 10
Code availability 10

**Foot and mouth disease (FMD)** 10
Unique combinations of author affiliation types 10
Collaboration types (aggregated) 10
Geographic connection of authors to the studied locations 12
Interventions 12
Modelling objectives 13
Outbreak types 13
Individual heterogeneity: agent-based versus compartmental models 13
Spatial heterogeneity 13
Model dynamics: deterministic vs stochastic 14
Outcomes measured 14
Parametrization methods 15
Validation methods 15
Sensitivity analysis 15
Data use and data availability 15
Code availability 16

#### Human vaccine-preventable diseases

##### Collaboration types (aggregated)

Table 1: Number of studies per collaboration type (Human VPDs)

| collab_type | n |
|---|---|
| purely_academic | 56.1% (128) |
| mixed | 43.9% (100) |
| Total | 100.0% (228) |

#### Locations studied

##### Real versus hypothetical

Table 2: Number of studies per location type

| location_type | n |
|---|---|
| real | 78.6% (195) |
| none | 21.4% (53) |
| Total | 100.0% (248) |

##### Larger than one country

Table 3: Number of studies per location larger than one country

| country_studied | n |
|---|---|
| west_africa | 47.4% (9) |
| global | 36.8% (7) |
| northern_hemisphere | 5.3% (1) |
| southeast_asia | 5.3% (1) |
| who_southeast_asia_region | 5.3% (1) |
| Total | 100.0% (19) |

##### Countries studied

Table 4: Number of studies per country

| country_studied | num_of_studies |
|---|---|
| US | 21.6% (38) |
| CN | 10.8% (19) |
| CA | 6.2% (11) |
| LR | 5.7% (10) |
| HK | 5.1% (9) |
| MX | 5.1% (9) |
| GB | 2.8% (5) |
| SL | 2.8% (5) |
| BR | 2.3% (4) |
| GN | 2.3% (4) |
| HT | 2.3% (4) |
| JP | 2.3% (4) |
| NL | 2.3% (4) |
| NO | 2.3% (4) |
| IL | 1.7% (3) |
| NG | 1.7% (3) |
| TW | 1.7% (3) |
| DK | 1.1% (2) |
| IN | 1.1% (2) |
| IT | 1.1% (2) |
| LY | 1.1% (2) |
| NE | 1.1% (2) |
| SG | 1.1% (2) |
| ZW | 1.1% (2) |
| AO | 0.6% (1) |
| AU | 0.6% (1) |
| BE | 0.6% (1) |
| BF | 0.6% (1) |
| CD | 0.6% (1) |
| CO | 0.6% (1) |
| CV | 0.6% (1) |
| ET | 0.6% (1) |
| FO | 0.6% (1) |
| FR | 0.6% (1) |
| GR | 0.6% (1) |
| HU | 0.6% (1) |
| IS | 0.6% (1) |
| NP | 0.6% (1) |
| PH | 0.6% (1) |
| PT | 0.6% (1) |
| SD | 0.6% (1) |
| SE | 0.6% (1) |
| TD | 0.6% (1) |
| TZ | 0.6% (1) |
| VE | 0.6% (1) |
| YE | 0.6% (1) |
| ZM | 0.6% (1) |
| Total | 100.0% (176) |

##### Continents studied

Table 5: Number of studies per continent

| continent_studied | num_of_studies |
|---|---|
| americas | 36.4% (68) |
| asia | 25.1% (47) |
| africa | 24.6% (46) |
| europe | 13.4% (25) |
| oceania | 0.5% (1) |
| Total | 100.0% (187) |

#### Geographic connection of authors to the studied locations

Table 6: Studies with at least one author affiliation in the location studied

| collab_type | country_studied | yes | no | Total |
|---|---|---|---|---|
| purely_academic | US | 94.4% (17) | 5.6% (1) | 18 |
| purely_academic | CN | 100.0% (8) | 0.0% (0) | 8 |
| purely_academic | HK | 77.8% (7) | 22.2% (2) | 9 |
| purely_academic | CA | 100.0% (3) | 0.0% (0) | 3 |
| purely_academic | TW | 100.0% (3) | 0.0% (0) | 3 |
| purely_academic | BR | 100.0% (2) | 0.0% (0) | 2 |
| purely_academic | MX | 66.7% (2) | 33.3% (1) | 3 |
| purely_academic | CO | 100.0% (1) | 0.0% (0) | 1 |
| purely_academic | GB | 50.0% (1) | 50.0% (1) | 2 |
| purely_academic | GR | 100.0% (1) | 0.0% (0) | 1 |
| purely_academic | HU | 100.0% (1) | 0.0% (0) | 1 |
| purely_academic | IL | 100.0% (1) | 0.0% (0) | 1 |
| purely_academic | IN | 50.0% (1) | 50.0% (1) | 2 |
| purely_academic | JP | 100.0% (1) | 0.0% (0) | 1 |
| purely_academic | NO | 100.0% (1) | 0.0% (0) | 1 |
| purely_academic | PH | 100.0% (1) | 0.0% (0) | 1 |
| purely_academic | PT | 100.0% (1) | 0.0% (0) | 1 |
| purely_academic | SE | 100.0% (1) | 0.0% (0) | 1 |
| purely_academic | SG | 100.0% (1) | 0.0% (0) | 1 |
| purely_academic | TZ | 100.0% (1) | 0.0% (0) | 1 |
| purely_academic | LR | 0.0% (0) | 100.0% (5) | 5 |
| purely_academic | HT | 0.0% (0) | 100.0% (3) | 3 |
| purely_academic | SL | 0.0% (0) | 100.0% (3) | 3 |
| purely_academic | GN | 0.0% (0) | 100.0% (2) | 2 |
| purely_academic | LY | 0.0% (0) | 100.0% (2) | 2 |
| purely_academic | ZW | 0.0% (0) | 100.0% (2) | 2 |
| purely_academic | AO | 0.0% (0) | 100.0% (1) | 1 |
| purely_academic | CD | 0.0% (0) | 100.0% (1) | 1 |
| purely_academic | CV | 0.0% (0) | 100.0% (1) | 1 |
| purely_academic | DK | 0.0% (0) | 100.0% (1) | 1 |
| purely_academic | ET | 0.0% (0) | 100.0% (1) | 1 |
| purely_academic | FO | 0.0% (0) | 100.0% (1) | 1 |
| purely_academic | NG | 0.0% (0) | 100.0% (1) | 1 |
| purely_academic | SD | 0.0% (0) | 100.0% (1) | 1 |
| mixed | US | 100.0% (19) | 0.0% (0) | 19 |
| mixed | CN | 100.0% (11) | 0.0% (0) | 11 |
| mixed | CA | 87.5% (7) | 12.5% (1) | 8 |
| mixed | MX | 83.3% (5) | 16.7% (1) | 6 |

Table 6: Studies with at least one author affiliation in the location studied *(continued)*

| collab_type | country_studied | yes | no | Total |
|---|---|---|---|---|
| mixed | NL | 100.0% (4) | 0.0% (0) | 4 |
| mixed | GB | 100.0% (3) | 0.0% (0) | 3 |
| mixed | JP | 100.0% (3) | 0.0% (0) | 3 |
| mixed | NO | 100.0% (3) | 0.0% (0) | 3 |
| mixed | BR | 100.0% (2) | 0.0% (0) | 2 |
| mixed | IL | 100.0% (2) | 0.0% (0) | 2 |
| mixed | IT | 100.0% (2) | 0.0% (0) | 2 |
| mixed | LR | 20.0% (1) | 80.0% (4) | 5 |
| mixed | AU | 100.0% (1) | 0.0% (0) | 1 |
| mixed | BE | 100.0% (1) | 0.0% (0) | 1 |
| mixed | BF | 100.0% (1) | 0.0% (0) | 1 |
| mixed | DK | 100.0% (1) | 0.0% (0) | 1 |
| mixed | FR | 100.0% (1) | 0.0% (0) | 1 |
| mixed | IS | 100.0% (1) | 0.0% (0) | 1 |
| mixed | NE | 50.0% (1) | 50.0% (1) | 2 |
| mixed | SG | 100.0% (1) | 0.0% (0) | 1 |
| mixed | TD | 100.0% (1) | 0.0% (0) | 1 |
| mixed | VE | 100.0% (1) | 0.0% (0) | 1 |
| mixed | GN | 0.0% (0) | 100.0% (2) | 2 |
| mixed | NG | 0.0% (0) | 100.0% (2) | 2 |
| mixed | SL | 0.0% (0) | 100.0% (2) | 2 |
| mixed | HT | 0.0% (0) | 100.0% (1) | 1 |
| mixed | NP | 0.0% (0) | 100.0% (1) | 1 |
| mixed | YE | 0.0% (0) | 100.0% (1) | 1 |
| mixed | ZM | 0.0% (0) | 100.0% (1) | 1 |
| Total | - | 72.6% (127) | 27.4% (48) | 175 |

##### Author affiliation in the studied location (overall)

Table 7: At least one author with an affiliation in the studied location

| collab_type | yes | no | Total |
|---|---|---|---|
| mixed | 83.1% (64) | 16.9% (13) | 77 |
| purely_academic | 67.5% (54) | 32.5% (26) | 80 |
| Total | 75.2% (118) | 24.8% (39) | 157 |

#### Interventions

##### Types of interventions

Table 8: Number of studies per intervention categories

| collab_type | no_vax | vax_combination_with_others | vax_single | Total |
|---|---|---|---|---|
| purely_academic | 53.1% (68) | 39.1% (50) | 7.8% (10) | 128 |
| mixed | 42.0% (42) | 45.0% (45) | 13.0% (13) | 100 |
| Total | 48.2% (110) | 41.7% (95) | 10.1% (23) | 228 |

##### Impact of vaccination

Table 9: Conclusions about impact of vaccination

| collab_type | yes | no | inconclusive | Total |
|---|---|---|---|---|
| mixed | 76.9% (10) | 15.4% (2) | 7.7% (1) | 13 |
| purely_academic | 90.0% (9) | 10.0% (1) | 0.0% (0) | 10 |
| Total | 82.6% (19) | 13.0% (3) | 4.3% (1) | 23 |

#### Modelling objectives

Table 10: Study objectives by collaboration type

| collab_type | both | future | past | Total |
|---|---|---|---|---|
| purely_academic | 3.1% (4) | 75.8% (97) | 21.1% (27) | 128 |
| mixed | 1.0% (1) | 70.0% (70) | 29.0% (29) | 100 |
| Total | 2.2% (5) | 73.2% (167) | 24.6% (56) | 228 |

#### Outbreak types

Table 11: outbreak types by collaboration type

| collab_type | hypothetical_outbreak | real_outbreak | Total |
|---|---|---|---|
| purely_academic | 52.3% (67) | 47.7% (61) | 128 |
| mixed | 47.0% (47) | 53.0% (53) | 100 |
| Total | 50.0% (114) | 50.0% (114) | 228 |

#### Individual heterogeneity: agent-based versus compartmental models

Table 12: How individuals were represented

| collab_type | compartments | agents | individuals_representation_other | Total |
|---|---|---|---|---|
| purely_academic | 80.5% (103) | 18.8% (24) | 0.8% (1) | 128 |
| mixed | 76.0% (76) | 22.0% (22) | 2.0% (2) | 100 |
| Total | 78.5% (179) | 20.2% (46) | 1.3% (3) | 228 |

#### Spatial heterogeneity

Table 13: Was space explicitly represented in the model

| collab_type | no | yes | Total |
|---|---|---|---|
| purely_academic | 73.4% (94) | 26.6% (34) | 128 |
| mixed | 68.0% (68) | 32.0% (32) | 100 |
| Total | 71.1% (162) | 28.9% (66) | 228 |

#### Model dynamics: deterministic vs stochastic

Table 14: Model dynamics (deterministic versus stochastic)

| collab_type | both | deterministic | stochastic | Total |
|---|---|---|---|---|
| purely_academic | 5.5% (7) | 69.5% (89) | 25.0% (32) | 128 |
| mixed | 7.0% (7) | 53.0% (53) | 40.0% (40) | 100 |
| Total | 6.1% (14) | 62.3% (142) | 31.6% (72) | 228 |

#### Outcomes measured

Table 15: Model outcomes by collaboration type

| collab_type | outcome_measured | num_of_studies |
|---|---|---|
| purely_academic | final_epidemic_size | 11.5% (54) |
| purely_academic | attack_rate | 6.2% (29) |
| purely_academic | timing_of_peak | 4.2% (20) |
| purely_academic | cost | 3.8% (18) |
| purely_academic | outbreak_duration_and_timing | 3.0% (14) |
| purely_academic | cases_averted | 2.5% (12) |
| purely_academic | intervention_coverage | 1.3% (6) |
| purely_academic | case_fatality | 1.1% (5) |
| purely_academic | cumulative incidence | 1.1% (5) |
| purely_academic | incidence | 1.1% (5) |
| purely_academic | control reproduction number | 0.8% (4) |
| purely_academic | peak magnitude | 0.8% (4) |
| purely_academic | peak magnitude | 0.6% (3) |
| purely_academic | campaign_duration | 0.6% (3) |
| purely_academic | deaths_averted | 0.6% (3) |
| purely_academic | hospitalizations | 0.6% (3) |
| purely_academic | basic reproduction number | 0.4% (2) |
| purely_academic | cumulative deaths | 0.4% (2) |
| purely_academic | basic reproduction number | 0.4% (2) |
| purely_academic | cumulative attack rate | 0.4% (2) |
| purely_academic | peak size | 0.4% (2) |
| purely_academic | qualys | 0.4% (2) |
| purely_academic | r0 | 0.4% (2) |
| purely_academic | total deaths | 0.4% (2) |
| purely_academic | and the individuals that have recovered and are immune to evd | 0.2% (1) |
| purely_academic | cumulative cases | 0.2% (1) |
| purely_academic | cumulative infections | 0.2% (1) |
| purely_academic | date of first reported cases | 0.2% (1) |
| purely_academic | epidemic prevention potential | 0.2% (1) |
| purely_academic | i and r curve | 0.2% (1) |
| purely_academic | incidence | 0.2% (1) |
| purely_academic | isolated or quarantined individuals | 0.2% (1) |
| purely_academic | net benefits | 0.2% (1) |
| purely_academic | number of latently infected individuals | 0.2% (1) |
| purely_academic | number of susceptible individuals | 0.2% (1) |
| purely_academic | number of times countermeasures are started | 0.2% (1) |
| purely_academic | peak day | 0.2% (1) |
| purely_academic | peak infections | 0.2% (1) |
| purely_academic | proportion of susceptible individuals | 0.2% (1) |
| purely_academic | proportion of time that infected size is above a threshold number of infecteds | 0.2% (1) |
| purely_academic | return on investment | 0.2% (1) |

Table 15: Model outcomes by collaboration type *(continued)*

| collab_type | outcome_measured | num_of_studies |
|---|---|---|
| purely_academic | risk of death | 0.2% (1) |
| purely_academic | transmission | 0.2% (1) |
| purely_academic | average overall effectiveness | 0.2% (1) |
| purely_academic | cumulative cases | 0.2% (1) |
| purely_academic | cumulative deaths | 0.2% (1) |
| purely_academic | cumulative hospital cases | 0.2% (1) |
| purely_academic | cumulative infections | 0.2% (1) |
| purely_academic | first arrival time | 0.2% (1) |
| purely_academic | force of infection | 0.2% (1) |
| purely_academic | funerals | 0.2% (1) |
| purely_academic | geometric mean number of infected hosts | 0.2% (1) |
| purely_academic | hospital notifications | 0.2% (1) |
| purely_academic | household reproduction number | 0.2% (1) |
| purely_academic | household reproduction number for | 0.2% (1) |
| purely_academic | incremental cost effectiveness ratio | 0.2% (1) |
| purely_academic | infection rate | 0.2% (1) |
| purely_academic | mortality | 0.2% (1) |
| purely_academic | mortality rate | 0.2% (1) |
| purely_academic | number of exposed and infectious individuals | 0.2% (1) |
| purely_academic | number of individuals in s | 0.2% (1) |
| purely_academic | number of simulations with epidemic outbreak | 0.2% (1) |
| purely_academic | peak daily infection | 0.2% (1) |
| purely_academic | peak incidence | 0.2% (1) |
| purely_academic | peak infection rate | 0.2% (1) |
| purely_academic | prevalence | 0.2% (1) |
| purely_academic | reproduction number | 0.2% (1) |
| purely_academic | total fraction of infected and exposed | 0.2% (1) |
| purely_academic | vaccine doses | 0.2% (1) |
| mixed | attack_rate | 6.6% (31) |
| mixed | final_epidemic_size | 6.6% (31) |
| mixed | cases_averted | 4.5% (21) |
| mixed | timing_of_peak | 3.4% (16) |
| mixed | outbreak_duration_and_timing | 2.8% (13) |
| mixed | hospitalizations | 1.9% (9) |
| mixed | intervention_coverage | 1.5% (7) |
| mixed | case_fatality | 1.1% (5) |
| mixed | cost | 1.1% (5) |
| mixed | cumulative incidence | 1.1% (5) |
| mixed | incidence | 1.1% (5) |
| mixed | r0 | 1.1% (5) |
| mixed | peak magnitude | 0.8% (4) |
| mixed | peak size | 0.6% (3) |
| mixed | qualys | 0.6% (3) |
| mixed | total deaths | 0.6% (3) |
| mixed | incidence | 0.4% (2) |
| mixed | population immunity | 0.4% (2) |
| mixed | cumulative attack rate | 0.4% (2) |
| mixed | cumulative cases | 0.4% (2) |
| mixed | deaths | 0.4% (2) |
| mixed | mortality rate | 0.4% (2) |
| mixed | peak incidence | 0.4% (2) |
| mixed | average expected number of cases | 0.2% (1) |
| mixed | average hospitalizations | 0.2% (1) |
| mixed | average infections | 0.2% (1) |
| mixed | average number of weeks lost | 0.2% (1) |
| mixed | case reproduction number | 0.2% (1) |

Table 15: Model outcomes by collaboration type *(continued)*

| collab_type | outcome_measured | num_of_studies |
|---|---|---|
| mixed | cumulative attack rate | 0.2% (1) |
| mixed | cumulative cases | 0.2% (1) |
| mixed | cumulative deaths | 0.2% (1) |
| mixed | incremental net benefits | 0.2% (1) |
| mixed | indirect protection | 0.2% (1) |
| mixed | maximum number of symptomatic cases per day | 0.2% (1) |
| mixed | number of courses of drug required to achieve containment | 0.2% (1) |
| mixed | total vaccinated | 0.2% (1) |
| mixed | vaccination coverage | 0.2% (1) |
| mixed | vaccine-derived virus prevalence | 0.2% (1) |
| mixed | years of life lost | 0.2% (1) |
| mixed | average deaths | 0.2% (1) |
| mixed | campaign_duration | 0.2% (1) |
| mixed | cumulative deaths | 0.2% (1) |
| mixed | cumulative infected | 0.2% (1) |
| mixed | dalys | 0.2% (1) |
| mixed | deaths_averted | 0.2% (1) |
| mixed | delay between epidemics | 0.2% (1) |
| mixed | duration of infection | 0.2% (1) |
| mixed | effective reproduction number | 0.2% (1) |
| mixed | effectiveness of vaccination strategies | 0.2% (1) |
| mixed | extra protective rate | 0.2% (1) |
| mixed | immunization threshold | 0.2% (1) |
| mixed | incremental cost effectiveness ratio | 0.2% (1) |
| mixed | incremental cost effectiveness ratio (icer) | 0.2% (1) |
| mixed | instantaneous reproduction number | 0.2% (1) |
| mixed | morbidity | 0.2% (1) |
| mixed | number of contacts traced | 0.2% (1) |
| mixed | number of deaths | 0.2% (1) |
| mixed | paralytic incidence | 0.2% (1) |
| mixed | peak difference | 0.2% (1) |
| mixed | peak prevalence | 0.2% (1) |
| mixed | prevalence of infection | 0.2% (1) |
| mixed | probability of preventing a large outbreak | 0.2% (1) |
| mixed | resistant cases | 0.2% (1) |
| mixed | risk of case importation | 0.2% (1) |
| mixed | time of detection | 0.2% (1) |
| Total | - | 100.0% (471) |

#### Sensitivity analysis

Table 16: Sensitivity analysis

| collab_type | no | yes | Total |
|---|---|---|---|
| purely_academic | 57.8% (74) | 42.2% (54) | 128 |
| mixed | 48.0% (48) | 52.0% (52) | 100 |
| Total | 53.5% (122) | 46.5% (106) | 228 |

##### Data use and data availability

Table 17: Data accessibility

| collab_type | yes | no | Total |
|---|---|---|---|
| purely_academic | 81.0% (51) | 19.0% (12) | 63 |
| mixed | 56.9% (37) | 43.1% (28) | 65 |
| Total | 68.8% (88) | 31.2% (40) | 128 |

##### Code availability

Table 18: Code availability

| collab_type | no | yes | Total |
|---|---|---|---|
| purely_academic | 98.4% (126) | 1.6% (2) | 128 |
| mixed | 98.0% (98) | 2.0% (2) | 100 |
| Total | 98.2% (224) | 1.8% (4) | 228 |

#### Foot and mouth disease (FMD)

##### Unique combinations of author affiliation types

Table 19: Number of studies by author affiliation type combination

| author_affiliation_type | n |
|---|---|
| academic_institutions | 44.0% (11) |
| academic_institutions + government_institutions | 24.0% (6) |
| government_institutions | 20.0% (5) |
| academic_institutions + government_institutions + NGO | 8.0% (2) |
| government_institutions + NGO | 4.0% (1) |
| Total | 100.0% (25) |

##### Collaboration types (aggregated)

Table 20: Number of studies per collaboration type

| collab_type | n |
|---|---|
| mixed | 56.0% (14) |
| purely_academic | 44.0% (11) |
| Total | 100.0% (25) |

**Proportions of the total publications per year by collaboration type**

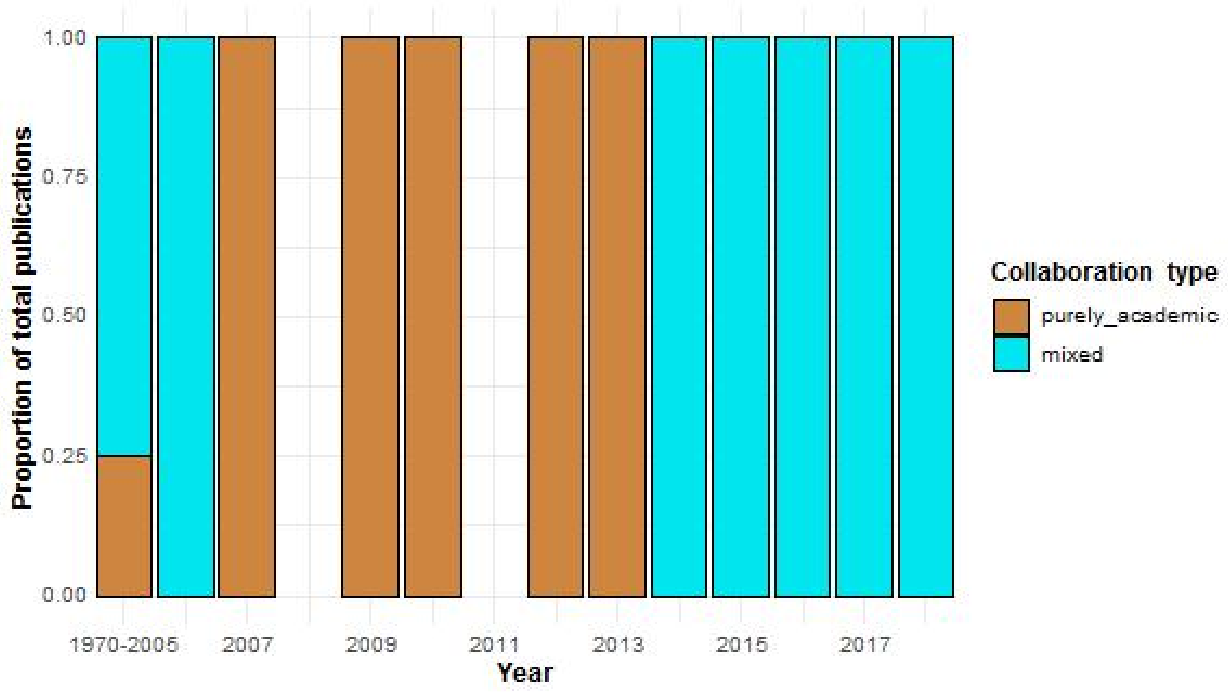


**Absolute number of publications per year by collaboration type**

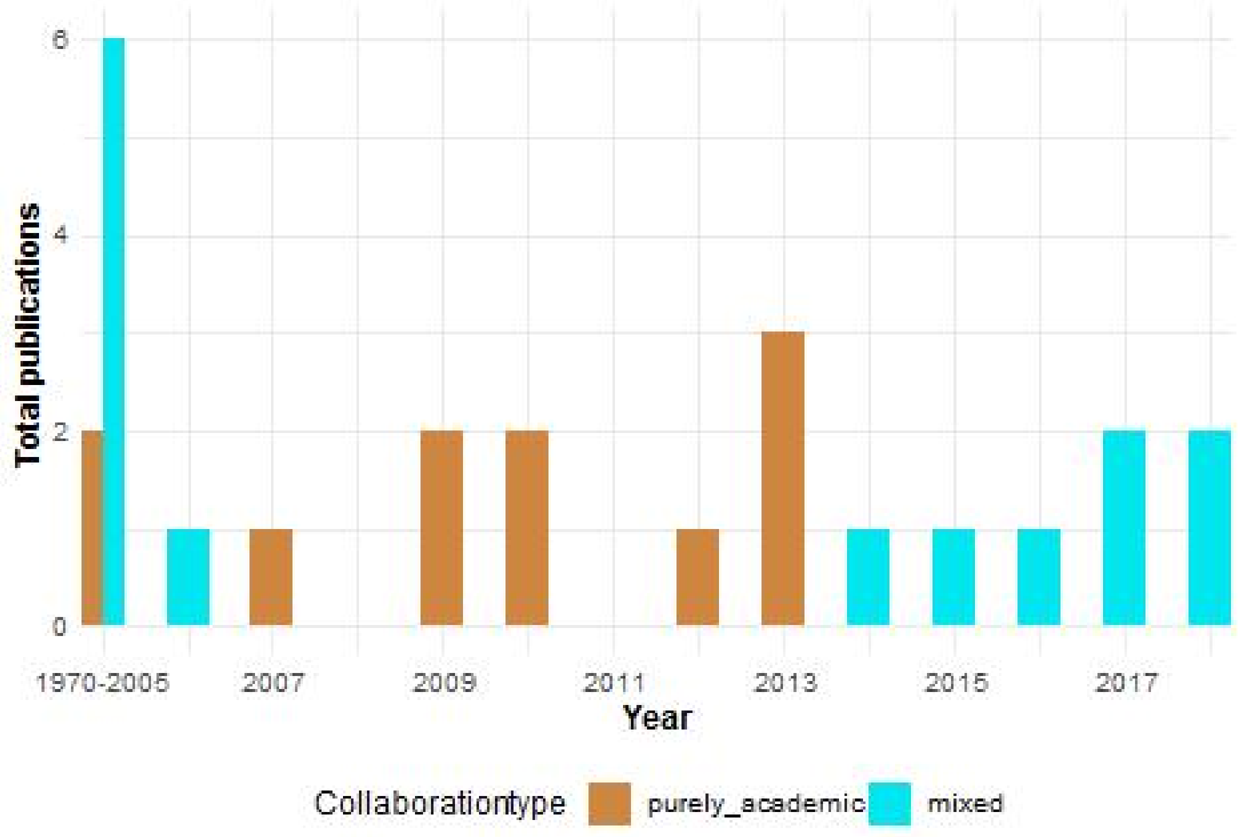

#### Geographic connection of authors to the studied locations

Table 21: Studies with at least one author affiliation in the location studied

| collab_type | country_studied | yes | no | Total |
|---|---|---|---|---|
| purely_academic | NL | 100.0% (4) | 0.0% (0) | 4 |
| purely_academic | GB | 66.7% (2) | 33.3% (1) | 3 |
| purely_academic | DK | 100.0% (2) | 0.0% (0) | 2 |
| purely_academic | US | 100.0% (1) | 0.0% (0) | 1 |
| purely_academic | ES | 100.0% (1) | 0.0% (0) | 1 |
| mixed | GB | 100.0% (2) | 0.0% (0) | 2 |
| mixed | FR | 100.0% (2) | 0.0% (0) | 2 |
| mixed | CH | 100.0% (2) | 0.0% (0) | 2 |
| mixed | US | 100.0% (1) | 0.0% (0) | 1 |
| mixed | NL | 100.0% (1) | 0.0% (0) | 1 |
| mixed | JP | 100.0% (1) | 0.0% (0) | 1 |
| mixed | AT | 100.0% (1) | 0.0% (0) | 1 |
| mixed | SE | 100.0% (1) | 0.0% (0) | 1 |
| mixed | AU | 100.0% (1) | 0.0% (0) | 1 |
| mixed | AU CA NZ GB US | 100.0% (1) | 0.0% (0) | 1 |
| mixed | UY | 0.0% (0) | 100.0% (1) | 1 |
| Total | - | 92.0% (23) | 8.0% (2) | 25 |

##### Aggregated

Table 22: Studies with at least one author affiliation in the location studied

| collab_type | yes | no | Total |
|---|---|---|---|
| mixed | 92.9% (13) | 7.1% (1) | 14 |
| purely_academic | 90.9% (10) | 9.1% (1) | 11 |
| Total | 92.0% (23) | 8.0% (2) | 25 |

#### Interventions

##### Types of interventions

Table 23: Number of studies per intervention type

| collab_type | vax_single | vax_combination_with_others | no_vax | Total |
|---|---|---|---|---|
| mixed | 71.4% (10) | 14.3% (2) | 14.3% (2) | 14 |
| purely_academic | 36.4% (4) | 45.5% (5) | 18.2% (2) | 11 |
| Total | 56.0% (14) | 28.0% (7) | 16.0% (4) | 25 |

**Impact of vaccination**

Table 24: Conclusions about impact of vaccination

| collab_type | no | inconclusive | yes | Total |
|---|---|---|---|---|
| mixed | 60.0% (6) | 20.0% (2) | 20.0% (2) | 10 |
| purely_academic | 50.0% (2) | 50.0% (2) | 0.0% (0) | 4 |
| Total | 57.1% (8) | 28.6% (4) | 14.3% (2) | 14 |

#### Modelling objectives

Table 25: Study objectives by collaboration type

| collab_type | future | past | Total |
|---|---|---|---|
| purely_academic | 63.6% (7) | 36.4% (4) | 11 |
| mixed | 71.4% (10) | 28.6% (4) | 14 |
| Total | 68.0% (17) | 32.0% (8) | 25 |

#### Outbreak types

Table 26: Study objectives by collaboration type

| collab_type | hypothetical_outbreak | real_outbreak | Total |
|---|---|---|---|
| purely_academic | 72.7% (8) | 27.3% (3) | 11 |
| mixed | 64.3% (9) | 35.7% (5) | 14 |
| Total | 68.0% (17) | 32.0% (8) | 25 |

#### Individual heterogeneity: agent-based versus compartmental models

Table 27: How individuals are represented

| collab_type | agents | compartments | Total |
|---|---|---|---|
| mixed | 71.4% (10) | 28.6% (4) | 14 |
| purely_academic | 81.8% (9) | 18.2% (2) | 11 |
| Total | 76.0% (19) | 24.0% (6) | 25 |

#### Spatial heterogeneity

Table 28: Models with spatial structure

| collab_type | yes | no | Total |
|---|---|---|---|
| purely_academic | 81.8% (9) | 18.2% (2) | 11 |
| mixed | 64.3% (9) | 35.7% (5) | 14 |
| Total | 72.0% (18) | 28.0% (7) | 25 |

#### Model dynamics: deterministic vs stochastic

Table 29: Model dynamics (deterministic versus stochastic)

| collab_type | deterministic | stochastic | Total |
|---|---|---|---|
| purely_academic | 36.4% (4) | 63.6% (7) | 11 |
| mixed | 50.0% (7) | 50.0% (7) | 14 |
| Total | 44.0% (11) | 56.0% (14) | 25 |

#### Outcomes measured

Table 30: Model outcomes by collaboration type

| collab_type | outcome_measured | num_of_studies |
|---|---|---|
| purely_academic | outbreak_duration_and_timing | 11.6% (8) |
| purely_academic | final_epidemic_size | 5.8% (4) |
| purely_academic | cost | 2.9% (2) |
| purely_academic | number of infected farms | 2.9% (2) |
| purely_academic | basic reproduction number | 1.4% (1) |
| purely_academic | export losses | 1.4% (1) |
| purely_academic | number of depopulated herds | 1.4% (1) |
| purely_academic | number of depopulated premises | 1.4% (1) |
| purely_academic | number of farms in control zones | 1.4% (1) |
| purely_academic | number of quarantined premises | 1.4% (1) |
| purely_academic | the number of pre-emptively slaughtered farms | 1.4% (1) |
| purely_academic | attack_rate | 1.4% (1) |
| purely_academic | effective reproduction number | 1.4% (1) |
| purely_academic | intervention_coverage | 1.4% (1) |
| purely_academic | lives_saved | 1.4% (1) |
| purely_academic | number of infected herds | 1.4% (1) |
| purely_academic | number of infected premises | 1.4% (1) |
| mixed | outbreak_duration_and_timing | 14.5% (10) |
| mixed | final_epidemic_size | 13.0% (9) |
| mixed | cost | 7.2% (5) |
| mixed | control costs | 1.4% (1) |
| mixed | employment effects | 1.4% (1) |
| mixed | income effects | 1.4% (1) |
| mixed | number of herds vaccinated | 1.4% (1) |
| mixed | number of premises culled | 1.4% (1) |
| mixed | output effects | 1.4% (1) |
| mixed | total number of herds culled | 1.4% (1) |
| mixed | total number of herds vaccinated | 1.4% (1) |
| mixed | compensation | 1.4% (1) |
| mixed | export losses | 1.4% (1) |
| mixed | intervention_coverage | 1.4% (1) |
| mixed | number of animals culled | 1.4% (1) |
| mixed | number of herds slaughtered | 1.4% (1) |
| mixed | total number of animals culled | 1.4% (1) |
| mixed | total number of herds surveilled | 1.4% (1) |

Table 30: Model outcomes by collaboration type *(continued)*

| collab_type | outcome_measured | num_of_studies |
| --- | --- | --- |
| mixed | total number of infected premises | 1.4% (1) |
| Total | - | 100.0% (69) |

#### Parametrization methods

Table 31: Model parametrization methods

| collab_type | literature and expert_opinion | literature and expert_opinion and fitted | fitted | literature | literature and fitted | expert_opinion | expert_opinion and fitted | Total |
| --- | --- | --- | --- | --- | --- | --- | --- | --- |
| mixed | 42.9% (6) | 7.1% (1) | 21.4% (3) | 21.4% (3) | 0.0% (0) | 7.1% (1) | 0.0% (0) | 14 |
| purely_academic | 9.1% (1) | 27.3% (3) | 18.2% (2) | 9.1% (1) | 18.2% (2) | 9.1% (1) | 9.1% (1) | 11 |
| Total | 28.0% (7) | 16.0% (4) | 20.0% (5) | 16.0% (4) | 8.0% (2) | 8.0% (2) | 4.0% (1) | 25 |

#### Validation methods

Table 32: Model validation methods

| collab_type | none | data | another_model | Total |
| --- | --- | --- | --- | --- |
| mixed | 71.4% (10) | 21.4% (3) | 7.1% (1) | 14 |
| purely_academic | 72.7% (8) | 27.3% (3) | 0.0% (0) | 11 |
| Total | 72.0% (18) | 24.0% (6) | 4.0% (1) | 25 |

#### Sensitivity analysis

Table 33: Sensitivity analysis

| collab_type | no | yes | Total |
| --- | --- | --- | --- |
| mixed | 71.4% (10) | 28.6% (4) | 14 |
| purely_academic | 36.4% (4) | 63.6% (7) | 11 |
| Total | 56.0% (14) | 44.0% (11) | 25 |

#### Data use and data availability

Table 34: Data accessibility

| collab_type | no | yes | Total |
| --- | --- | --- | --- |
| mixed | 100.0% (6) | 0.0% (0) | 6 |
| purely_academic | 62.5% (5) | 37.5% (3) | 8 |
| Total | 78.6% (11) | 21.4% (3) | 14 |

#### Code availability

Table 35: Code availability

| collab_type | no |
|---|---|
| purely_academic | 44.0% (11) |
| mixed | 56.0% (14) |
| Total | 100.0% (25) |
